# Genetic analysis of selection bias in a natural experiment: Investigating in-utero famine effects on elevated body mass index in the Dutch Hunger Winter Families Study

**DOI:** 10.1101/2023.10.23.23297381

**Authors:** Jiayi Zhou, Claire E. Indik, Thomas B. Kuipers, Chihua Li, Michel G. Nivard, Calen P. Ryan, Elliot M. Tucker-Drob, M. Jazmin Taeubert, Shuang Wang, Tian Wang, Dalton Conley, Bastiaan T. Heijmans, LH Lumey, Daniel W. Belsky

## Abstract

Natural-experiment designs that compare survivors of in-utero famine exposure to unaffected controls suggest that in-utero undernutrition predisposes to development of obesity. However, birth rates drop dramatically during famines. Selection bias could arise if factors that contribute to obesity also protect fertility and/or fetal survival under famine conditions. We investigated this hypothesis using genetic analysis of a famine-exposed birth cohort. We genotyped participants in the Dutch Hunger Winter Families Study (DHWFS, N=950; 45% male), of whom 51% were exposed to the 1944-1945 Dutch Famine during gestation and 49% were their unexposed same-sex siblings or “time controls” born before or after the famine in the same hospitals. We computed body-mass index (BMI) polygenic indices (PGIs) in DHWFS participants and compared BMI PGIs between famine-exposed and control groups. Participants with higher polygenic risk had higher BMIs (Pearson r=0.42, p<0.001). However, differences between BMI PGIs of famine-exposed participants and controls were small and not statistically different from zero across specifications (Cohen’s d=0.10, p>0.092). Our findings did not indicate selection bias, supporting the validity of the natural-experiment design within DHWFS. In summary, our study outlines a novel approach to explore the presence of selection bias in famine and other natural experiment studies.

## INTRODUCTION

The roots of age-related chronic disease extend backward in development to the earliest stages of life. The dominant conceptual model linking early adverse exposures to later-life health and aging is the Developmental Origins of Health and Disease (DOHaD) hypothesis.^1^ The most basic formulation of the DOHaD hypothesis posits that adverse early exposures sculpt development in ways that predispose to a range of later-life health problems, including through epigenetic mechanisms.^2–4^ An alternative conceptual model, broadly termed the in-utero-selection hypothesis, suggests that adverse exposures can influence fertility and/or fetal mortality in ways that change characteristics of the offspring generation.^5^

A canonical DOHaD process is the programming of obesity risk by in-utero undernutrition. In animal experiments, dietary restriction of pregnant females induces vulnerability to obesity in their offspring.^1^ Similarly, in human observational studies, babies with low birth weight and other indications of in-utero undernutrition more often grow up to develop obesity.^6^ While experimental manipulation of early-life nutrition in humans is unethical, researchers have pursued quasi-experimental tests of DOHaD through follow-up of cohorts exposed to famine in-utero.^7,8^ Such “natural experiments” can identify causal effects of in-utero undernutrition, especially if the population at risk, the timing and degree of exposure, and relevant health outcomes can be accurately defined.^9^ In well-designed famine-natural-experiment studies, compared with unexposed controls, famine-exposed births more often develop obesity as adults.^10–12^

An assumption in famine-natural-experiment studies is that famine survivors and controls are “exchangeable,” i.e. they differ only in their exposure to the famine. However, famine is associated with a dramatic reduction in births. For example, in the case of the Dutch Famine of 1944-45, the birth rate fell by as much as 50%.^13^ In this context, famine survivors may not be exchangeable with controls. Under the in-utero-selection hypothesis, there are at least two paths through which the extreme winnowing of births under famine conditions could result in differences in obesity risk between famine survivors and controls. The first path is that adults who carry higher genetic risk for obesity may more often maintain their fertility under famine conditions, e.g., because they carry a larger buffer against undernutrition-induced infertility.^14^ The second path is that fetuses that inherit higher genetic risk for obesity may more often survive gestation under famine conditions, e.g., because of a thriftier physiology.^15^ In summary, the same traits that predispose to overweight under non-famine conditions could protect parental health and fertility and/or fetal survival under conditions of famine.

There is evidence to support the in-utero-selection hypothesis from epigenetic analysis of DNA methylation from famine survivors and controls.^16^ Here we investigate the hypothesis using genetic analysis to compare survivors of in-utero famine with control individuals born before and after the famine in the same location. Genotypes are fixed at conception and therefore cannot be modified by famine exposure. Genetic factors are also powerful determinants of risk for obesity; studies of twins and families estimate upwards of two thirds of variation in general metabolic outcomes is attributable to genetic factors.^17^ If the selection hypothesis is correct, famine-exposed individuals should carry higher genetic risk as compared to the unexposed comparison group. Genome-wide association studies (GWAS) have identified hundreds of common variants for elevated body mass index.^18,19^ Our study design utilizes risk scores (referred to as “polygenic indices”) composed of genetic variants discovered in these GWAS to test whether survivors of in-utero famine carry elevated genetic risk for obesity.

We analyzed data from the Dutch Hunger Winter Family Study (DHWFS), a landmark birth cohort study of survivors of in-utero famine, matched controls born in the same hospitals before and after the famine, and siblings of these individuals.^20^ Prior studies of this cohort establish that famine survivors exhibit excess risk of obesity.^10^ In the current study, we analyzed DNA samples from the cohort members to compute polygenic indices quantifying inherited genetic risk for obesity. We compared famine-survivors with unexposed participants to test the selection hypothesis. Given the numbers of exposed and unexposed participants in DHWFS, our analysis has >80% power to detect differences between groups d>0.18. Assuming a polygenic index effect-size of r=0.3^19^ and a BMI difference between exposed and unexposed participants of d=0.4,^10^ a polygenic index difference between exposed and unexposed groups of d=1.3 (0.4/0.3) would be required to explain the observed famine effect. Our study is therefore well powered to detect the presence of selection bias that could confound even a fraction of the observed BMI difference between famine-exposed and control participants.

## METHOD

### Study setting: The Dutch Hunger Winter of 1944-1945

The Dutch famine occurred because of a food-supply embargo by German occupying forces in early October 1944. The severity and widespread nature of the famine are well documented.^13,20,21^ Before the famine, nutrition in the Netherlands had been adequate. Official rations, which eventually consisted of little more than bread and potatoes, fell below 900 kcal/day in late November 1944, and were as low as 500 kcal/day by April 1945. The macronutrient composition of the ration remained relatively stable over this period, but the composition of non-ration foods changed, with a reduction in fat intake. The famine ceased with liberation in May 1945, after which Allied food supplies were distributed.

### Participants

Famine-exposed individuals were identified from review of archival obstetric records. We selected all the 2,417 singleton births between 1 February 1945 and 31 March 1946 at three institutions in famine-exposed cities in the western Netherlands whose mothers were exposed to the famine during or immediately preceding that pregnancy. Time Controls were selected from births at the same hospitals as the famine-exposed group during 1943 and 1947 (2 years before and 2 years after the famine). We sampled an equal number of births each month, allocated across the three institutions according to their size, to obtain 890 singleton births.

Of the total famine-exposed and time-control births, current addresses were able to be traced for 70%. These individuals were invited by mail to join the study and were additionally asked if a same-sex sibling born before or after the famine would be available to participate. A total of n=547 of the famine-exposed group, n=176 of the time-control group, and n=308 same-sex unexposed siblings consented to participate and underwent a computer-assisted structured interview by telephone. Comparing traced and untraced individuals, or respondents and non-respondents to the invitation letter, or actual participants in the study vs non-participants) there were no clinical differences in their characteristics at birth (birth weight, length, placental weight, maternal age or birth order).^20^

Data Collection was conducted in 2003-2005, approximately six decades after the famine. Of the n=1,031 participants who were interviewed, n=971 also participated in a clinic exam. Following the Helsinki guidelines, we obtained ethical approval both from the Institutional Review Board of Columbia University Medical Center and from the Medical Ethical Committee of the Leiden University Medical Center (LUMC). The study participants provided verbal consent in a telephone interview, and in case of clinical examinations, a written informed consent was obtained.

The analysis sample for this study was formed from participants in the clinic exam who provided a blood sample from which DNA was extracted and stored at LUMC. After quality controls, genetic datasets were available for n=950. These individuals formed our analysis sample (**Figure 1**).

**Figure 1.**
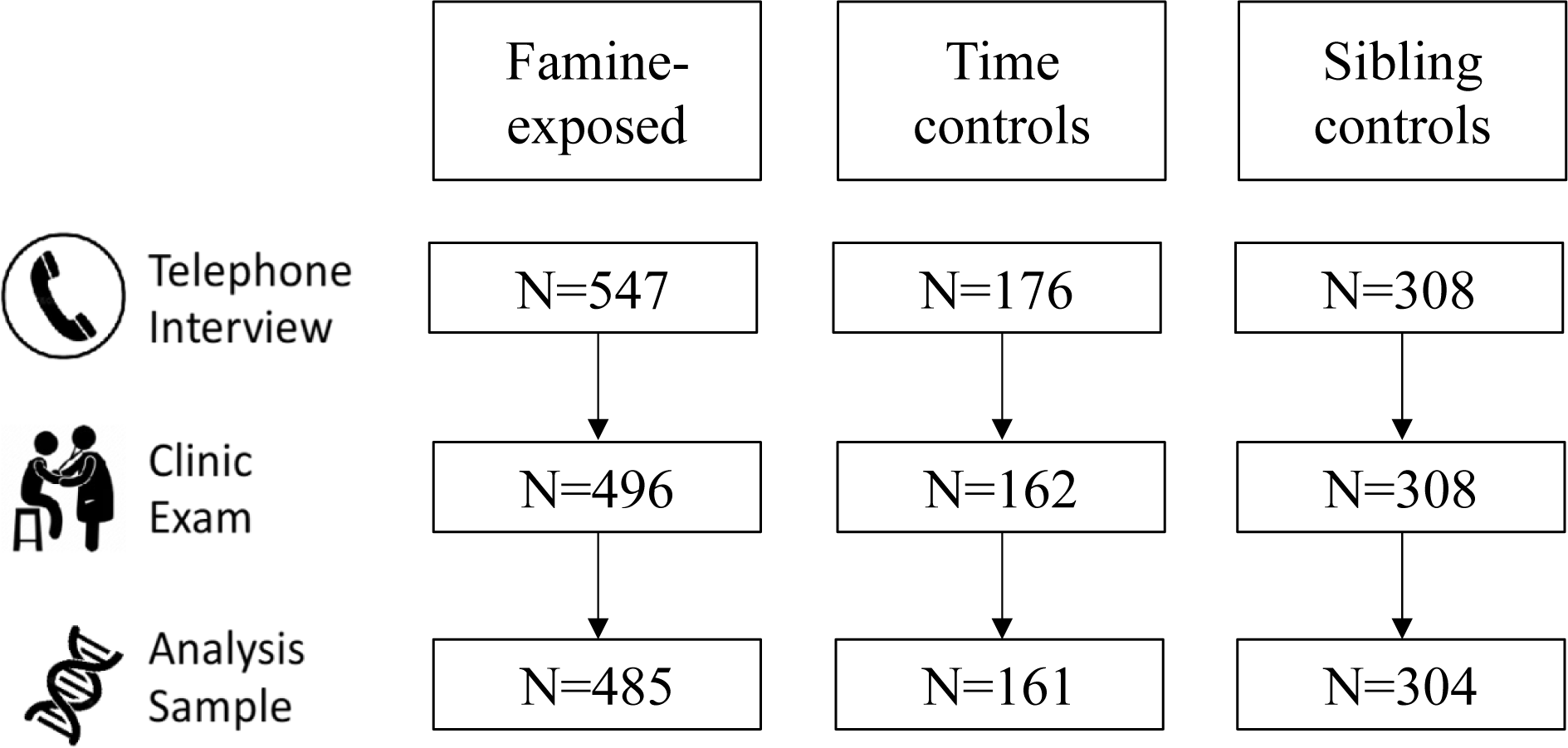
Flow diagram of the Dutch Hunger Winter Families Study. The figure shows how the analysis sample size was arrived at in each step for survivors of in-utero famine exposure, time controls, and same-sex sibling controls. N=1,031 participants completed telephone interviews. Of this group, 971 participated in the clinic exam. Available DNA extracted from these blood samples allowed us to generate genetic data were generated for 950 of these individuals. The figure illustrates the number of individuals in each exposure and control group included in the telephone interview, clinic examination, and analysis sample.

### Measures

#### Famine exposure

We defined the period of famine from archival records of weekly ration distributions, as described previously.^16,20,22,23^ Briefly, the start of the famine-exposure period was defined as November 26, 1944, based on the threshold <900 kcal/day of distributed food rations. The end of the famine-exposure period was defined as May 12, 1945, one week following the German surrender. Participants’ exposure during gestation was determined from the date of their mother’s last menstrual period (LMP) and their date of birth. In cases where the LMP date was missing or implausible (12% of births), LMP was estimated from birth-record data on birth weight and date of birth using tables of gender, parity, and birth weight specific gestational ages from the combined birth records of the Amsterdam Midwives School (1948-57) and the University of Amsterdam Wilhelmina Gasthuis Hospital (1931-65).

Famine-exposed participants experienced an average of 17 weeks of gestation during which ration distributions were <900kcal/day. Following prior work with the cohort,^20,23–25^ participants were classified as famine exposed during each of four 10-week gestational periods on the basis of ration distributions. For each individual, average rations were calculated for each 10-week period of gestation and periods with average rations <900 kcal/day were classified as famine-exposed. Among the n=547 participants recruited as famine exposed, n=403 met criteria for exposure in at least one 10-week gestational period. A further n=82 had LMP dates prior to the end of the famine, but average ration distributions during their first 10 weeks of gestation <900kcal/day. A final n=62 had LMP dates after the end of the famine. Gestational periods for famine-exposed participants and time-controls are shown in **Figure 2** and **Supplemental Table 2.**

**Figure 2.**
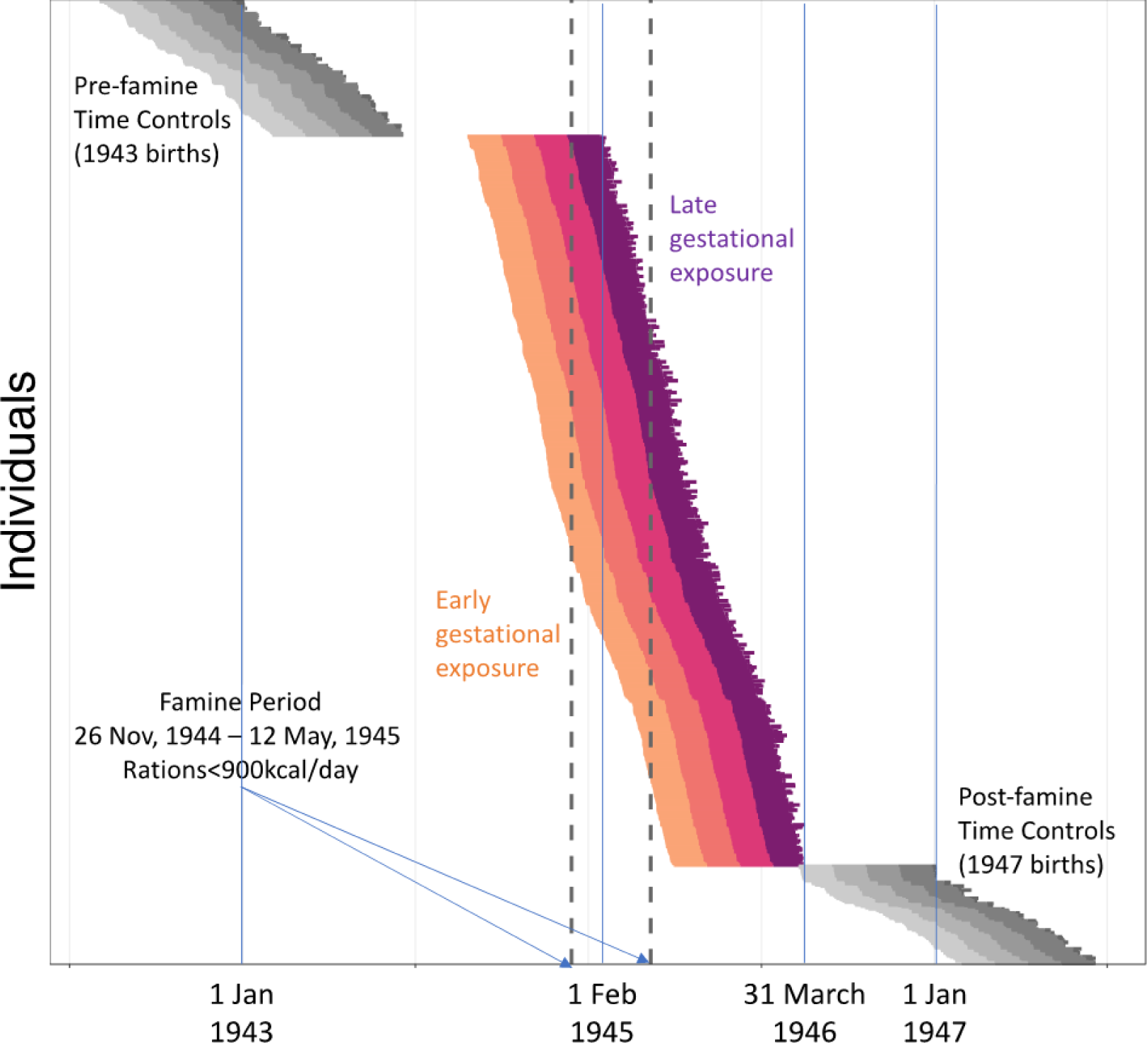
Gestational timing of exposure to famine in the Dutch Hunger Winter Families Study. The figure shows individual gestations of N=547 famine-exposed participants (colored lines) and N=176 time controls (gray lines). Each gestation is plotted as a single horizontal line. The start of the line is the date of the mother’s last menstrual period (LMP). The end of the line is the participant’s date of birth. Individual gestations are plotted from the top of the graph to the bottom, ordered by LMP date. For the famine-exposed participants, the segment of each line showing the first 10 weeks of gestation is colored gold. The segment showing the second 10 weeks is colored orange. The segment showing the third 10 weeks is colored red. The segment showing the last 10 weeks is colored purple. For the time controls, 10-week gestational periods are colored in gray, with lighter shades for the earlier gestational periods. The x-axis shows the date. The vertical dashed lines show the start and end of the famine exposure period (November 26, 1944 - May 12, 1945).

**Table 1.** Sample Characteristics. The table shows the characteristics of the analysis sample (N = 950) included from the Dutch hunger winter family cohort with genetic data. The analysis sample was separated into three categories based on exposure status. The data in the table show that the age and sex ratio of participants in our analysis sample are comparable across the three exposure categories. Body mass index (BMI) were measured in the sample participating the clinical exam.

| Analysis Sample (N = 950) |  |  |  |  |  |  |  |  |  |  |  |  |
| --- | --- | --- | --- | --- | --- | --- | --- | --- | --- | --- | --- | --- |
|  | Famine-exposed<br>(N = 485) |  |  |  | Time Controls<br>(N = 161) |  |  |  | Sibling Controls<br>(N = 304) |  |  |  |
|  | N | (%) | Mean | SD<sup>1</sup> | N | (%) | Mean | SD | N | (%) | Mean | SD |
| Age at Clinical Exam |  |  | 58.8 | 0.5 |  |  | 59.2 | 2.0 |  |  | 57.0 | 6.0 |
| Male | 299 | (47%) |  |  | 72 | (45%) |  |  | 129 | (42%) |  |  |
| BMI (N = 947) | 483 |  | 28.3 | 4.8 | 161 |  | 27.1 | 4.2 | 303 |  | 27.0 | 4.2 |
| <sup>1</sup> SD: Standard deviation |  |  |  |  |  |  |  |  |  |  |  |  |

<sup>1</sup> SD: Standard deviation

#### Body Mass Index

Body mass index (BMI) was computed from measurements of weight measured in light clothes with a SECA scale (SECA, Hamburg, Germany) and height, measured with a fixed stadiometer. BMI was calculated as weight in kilograms divided by height in meters squared (kg/m2).

#### Polygenic Index

Genotyping and QC were performed by the Human Genomics Facility at the Erasmus University Medical Center (Rotterdam, Netherlands) from DNA extracted from whole blood using the Infinium™ Global Screening Array (version-24 v3.0 Bead-Chip arrays; Illumina). SNPs with call rates <97.5% or not in Hardy-Weinberg equilibrium (p>1×10^−4^) were removed. The final dataset was imputed to the 1000 Genomes Phase-3 version-5 reference panel^26–28^ using Minimac4 (https://genome.sph.umich.edu/wiki/Minimac4). The resulting genetic dataset includes n=950 participants.

We measured participants’ genetic risk for obesity using a polygenic index (PGI). PGIs summarize genome-wide genetic influence on a phenotype based on genome-wide association study (GWAS) results.^29^ PGIs are calculated as the average weighted count of phenotype-associated alleles across all SNPs included in the GWAS, where the weights are derived from GWAS results. We computed our PGI using the GIANT Consortium’s most recent BMI GWAS^30^ according to the method of Khera et al.^19^. The final BMI PGI was computed with LDpred2’s ‘auto’ model.^31^ Details are in the **Supplemental Methods**.

DHWFS is homogenous with respect to genetic ancestry and migration status; all but a handful women were white and born in the Netherlands and 95% of their fathers were born in the Netherlands. To account for residual population stratification, we computed principal components from the genome-wide SNP matrix ^32,33^ using the “PC-AiR” method from the GENESIS Bioconductor library in R, which accommodates data that include genetic relatives.^34,35^ For analysis, we residualized the polygenic index for the first 10 principal components and standardized residual values to mean M=0, standard deviation SD=1 in DHWFS.^36^

### Statistical analysis

We analyzed continuous outcomes using linear regression. Models included covariates for age, age squared, and sex. To account for DHWFS’s sibling structure, we used generalized estimating equations (GEE).^37^ We conducted analysis of within-family sibling differences using econometric fixed-effects.^38^

We tested associations of famine exposure and genetic risk with BMI using linear regression. We regressed BMI on each of the risk factors in separate regressions (i.e., one risk factor per model). Models included covariates for age, age squared, and sex. We tested gene-environment correlation (rGE) by regressing the polygenic index measures of genetic risk of BMI on famine exposure and the same covariates included in the main-effect regressions. The rGE association was tested by the regression coefficient of famine exposure. We tested gene-environment interaction (GxE) on BMI by regressing BMI on famine exposure, the polygenic index, a cross-product term of famine exposure and PGI for their interaction, and the same panel of covariates as the main-effect and rGE models. We tested the GxE hypothesis by testing the regression coefficient of the cross-product term. We report effect sizes in standard deviation units. We refer to effect estimates for association with dichotomous exposure variables as Cohen’s d (standard deviation difference between exposed and control). We refer to effect estimates for associations with continuous exposure variables as Pearson r (standard deviation difference in outcome per standard deviation difference in exposure).

All analyses were conducted with R (version 4.1.2). GEE analysis was performed with ‘geeglm’ function from ‘geepack’ libary (version 1.3.9).^39^ Sibling comparison analysis was performed with ‘plm’ function from ‘plm’ library (version 2.6-2).^40^

## RESULTS

We conducted genetic analysis of associations between in-utero famine exposure and elevated body-mass index (BMI). Our analysis proceeded in three steps. First, we tested differences in BMI between famine-exposed participants and controls. Next, we tested for gene-environment correlation (rGE), i.e. differences in genetic risk (PGI) for higher BMI between famine-exposed participants and controls. We then integrated these analyses by re-estimating famine-exposure associations with BMI including covariate adjustment for genetic risk. Finally, we tested for gene-environment interaction (GxE), i.e. differential association of genetic risk with BMI between exposed participants and controls. In each of these analyses, we compared famine-exposed participants to three control groups to establish the robustness of differences. The first comparison group included all control participants. The second comparison group included the subgroup of control participants recruited as time controls. The third comparison group included the subgroup of control participants who were unexposed siblings of the famine-exposed group; this comparison group was used to conduct sibling difference analyses. The purpose of the sibling-difference analysis was to evaluate whether associations might be confounded by factors common to siblings within a family, such as shared childhood-household conditions.

Survivors of in-utero famine exposure had higher BMI six decades later when compared with unexposed controls. Effect estimates were similar across the three comparison groups (full DHWFS comparison Beta=0.28, 95%CI [0.16, 0.40], p<0.001; exposed vs. time controls Beta=0.25, 95%CI [0.06, 0.43], p=0.009; sibling difference Beta=0.29, 95%CI [0.12-0.46], p<0.001).

Before conducting the test of gene-environment correlation, we confirmed BMI PGI within DHWFS. Participants with higher PGI-measured genetic risk had higher BMIs. Effect estimates were similar across comparison groups (full-DHWFS-comparison Beta=0.42, 95% CI [0.35-0.49], p<0.001; exposed-vs.-time-control Beta=0.41, 95% CI [0.32, 0.49], p<0.001; sibling-difference Beta=0.46, 95% CI [0.31-0.61], p<0.001). However, we did not observe strong evidence of rGE. BMI PGIs were somewhat higher for the famine-exposed group as compared with controls, but differences were small and not statistically different from zero (full-DHWFS-comparison Beta=0.10, 95% CI [-0.02-0.21], p=0.092; exposed-vs.-time-control Beta=0.04, 95% CI [-0.14, 0.22], p=0.674; sibling-difference Beta=0.04, 95% CI [-0.11, 0.18], p=0.601).

We next tested inclusion of the BMI PGI as a covariate in analysis of famine effects on BMI. If famine-associated selection on genetic risk for BMI induced a spurious association, then covariate adjustment for the PGI should attenuate famine exposure effect-sizes. However, the effect-size in our adjusted model was similar to the effect-size in our original model. Effect-sizes comparisons are shown in **Figure 3**.

**Figure 3.**
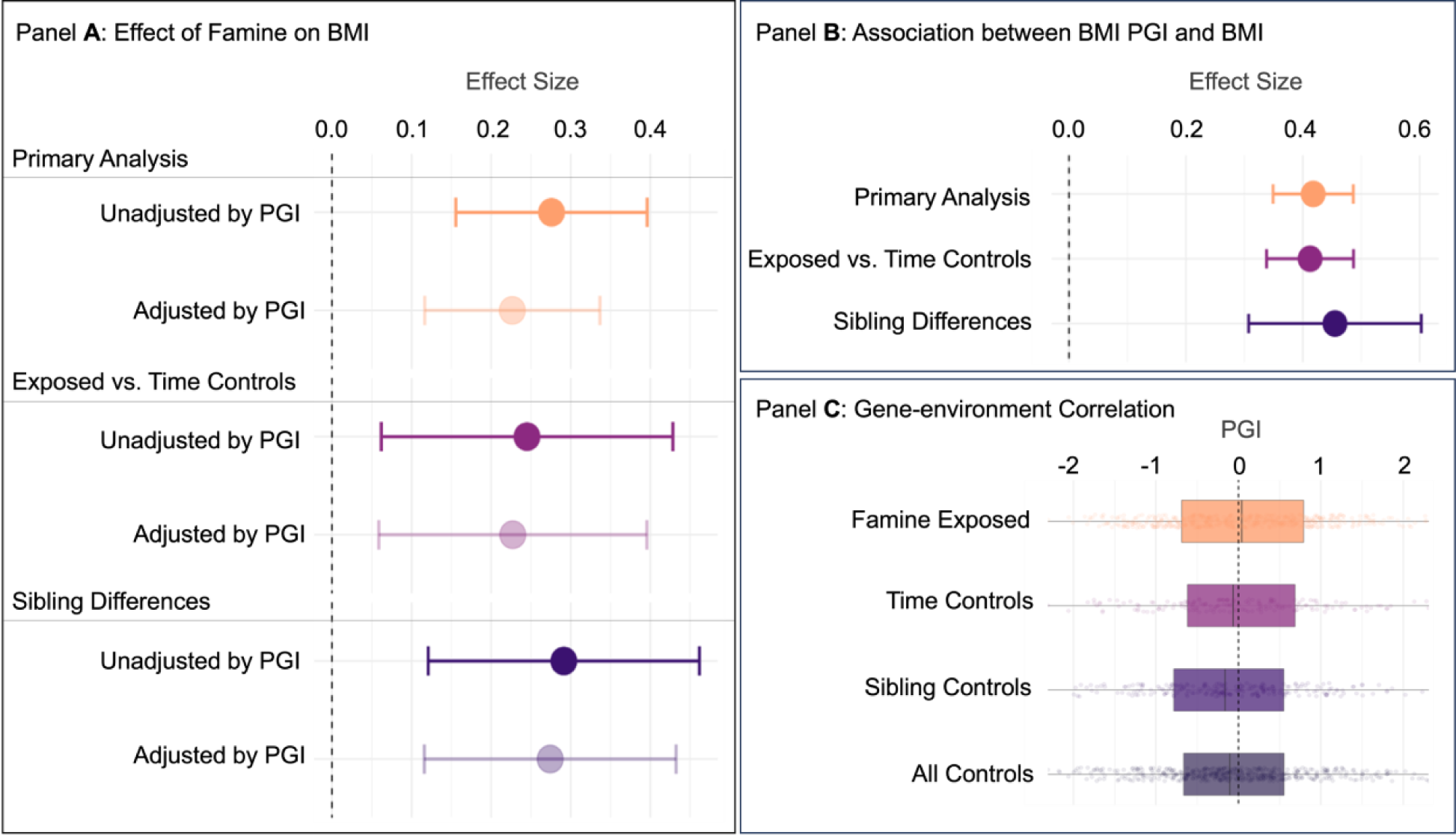
Main effect of famine with and without adjustment for body mass index (BMI) polygenic index (PGI). **Panel A** shows main effects of famine exposure on BMI. Standardized effect size is on the x-axis, and model type is shown on the y-axis. Models were grouped into three classes: “Primary analysis”, “Sibling Difference”, and “Exposed vs. Time Controls”, where each group contains two models: the “Unadjusted by PGI” model and the “Adjusted by PGI” model which included an additional BMI PGI covariate. In the “Primary Analysis” group, both “Unadjusted by PGI” and “Adjusted by PGI” models included all participants in our analysis sample with available data on BMI (n=947) and was fitted using linear regression within a generalized estimating equations (GEE) framework to account for non-independence of data from sibling pairs. Models included covariates for participant age, age-squared, and sex. In the “Exposed vs. Time Controls” group, both “Unadjusted by PGI” and “Adjusted by PGI” models included the subset of the Primary Analysis sample consisting of famine exposure participants and their time controls (n= 644) and was fitted using linear regression within a generalized linear model (GLM) with a sex covariate. In the “Sibling Difference” group, both “Unadjusted by PGI” and “Adjusted by PGI” models included the subset of the Primary Analysis sample consisting of sibling pairs discordant for famine exposure (n=226 pairs) and was fitted using fixed effects regression. Models included covariates for age and age-squared (all sibling pairs were of the same sex). For all models, standardized effect sizes were shown as a dot representing the regression beta coefficient of famine with an error bar indicating the 95% confidence intervals (CI). **Panel B** shows the association between BMI PGI and BMI. Standardized effect size is on the x-axis, and model type is shown on the y-axis. In the “Primary Analysis” group, BMI was regressed on BMI PGI using linear regression within a generalized estimating equations (GEE) model including all participants in our analysis sample with available data on BMI (n=947). Models included covariates for participant age, age-squared, and sex. The “Exposed vs. Time Controls” group model included the subset of the Primary Analysis sample consisting of famine exposure participants and their time controls (n= 644) and was fitted using linear regression within a generalized linear model (GLM) with a sex covariate. The “Sibling Difference” model included the subset of the Primary Analysis sample consisting of sibling pairs discordant for famine exposure (n=226 pairs) and was fitted using fixed effects regression. Models included covariates for age and age-squared (all sibling pairs were of the same sex). For all models, standardized effect sizes were shown as a dot representing the regression beta coefficient of BMI PGI with an error bar indicating the 95% confidence intervals (CI).**Panel C** presents a set of box plots reporting the distribution of BMI PRS by famine exposure categories: “Famine-exposed” (n = 485), “Time Controls” (n = 161), “Sibling Controls” of famine-exposed (simplified as “Sibling Controls” in the plot legend: n = 226), and “All Controls” consist of time controls, sibling controls of time controls, and sibling controls of famine-exposed (n = 465)

Our final analysis tested for GxE. In stratified analysis, effect-sizes for PGI associations with BMI were somewhat larger for the famine-exposed group as compared with controls (effect of PGI on BMI among famine-exposed Beta=0.43, 95% CI [0.34, 0.52], p<0.001; effect of PGI in controls Beta=0.38, 95% CI [0.30-0.47], p<0.001; effect of PGI among time controls Beta=0.33, 95% CI [0.19, 0.46], p<0.001; effect of PGI among sibling controls of famine-exposed Beta=0.39, 95%CI [0.28, 0.50], p<0.001). However, models testing GxE found that differences were not statistically different from zero (GxE p>0.457). Genetic associations with BMI in each exposure and control group are shown in **Figure 4**. Full results from all regression analysis are reported in **Table S4.**

**Figure 4.**
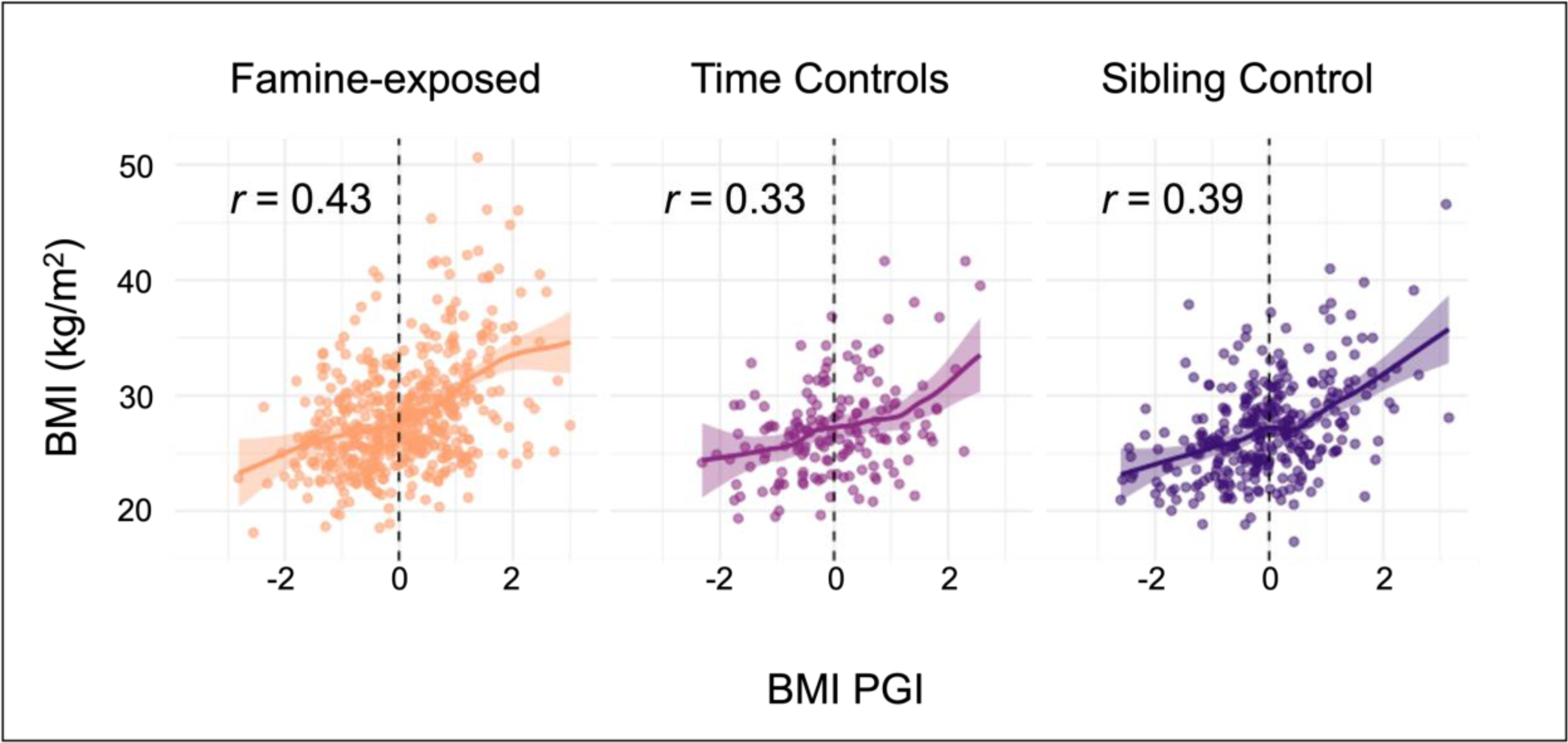
Body mass index PGI associations with body mass index in survivors of in-utero famine, time-controls, and sibling controls in the Dutch Hunger Winter Families Study. The figure shows scatterplots of associations between the body-mass index (BMI) polygenic index (PGI) and BMI in three groups of participants from the Dutch Hunger Winter Families Study (DHWFS). The X-axis shows participants’ polygenic indices on a standardized scale (M=0, SD=1). The Y-axis shows their raw BMIs with the unit (kg/m2). Standardized coefficients (interpretable as Pearson r) from generalized-linear model analysis are reported for each group. The figure shows that associations of the BMI PGI with BMI are similar across the three groups.

We conducted sensitivity analyses to evaluate the impact of the duration and gestational timing of famine exposure, of sex differences, of alternative methods of polygenic index construction, and to evaluate the impact of measurement error in the polygenic index. These are described in the **Supplemental Materials**. Results were consistent with our primary analysis and are reported in **Supplemental Tables S5-S17**, and **Supplemental Figures S1-4**.

## DISCUSSION

We conducted genetic analysis of the effect of in-utero famine exposure on midlife body-mass index (BMI) in the Dutch Hunger Winter Families Study (DHWFS) to evaluate potential confounding of the famine natural experiment by selective fertility and/or fetal survival. Survivors of in-utero famine exposure tend to show elevated BMI as compared with controls.^11,12,41^ Our analysis tested if this difference could be confounded by a tendency of famine-survivors to carry high genetic risk of obesity. Results did not support the presence of substantial confounding of in-utero famine effects by genetic risk. Among participants in DHWFS, both famine exposure and genetic risk were associated with higher BMI. However, genetic risk for obesity, as measured by a BMI polygenic index, was similar among survivors of in-utero famine exposure and controls. These findings support the validity of the natural-experiment design of the Dutch Hunger Winter Families Study and more generally suggest that birth cohorts of famine-survivors and matched time-controls are not confounded by large-effect genetic differences related to the risk of obesity.

DHWFS is a relatively small cohort by the standards of genetics research. The BMI PGI we used, while a powerful predictor of BMI and obesity risk, ^19^ is an incomplete summary of genetic risks for obesity.^42,43^ Therefore, our analysis cannot rule out the possibility that famine survivors carry some excess genetic risk relative to the general population. However, our findings establish that any such differences are likely to be of small effect and therefore, in the case of obesity, unlikely to confound the general conclusions of famine studies that lack genetic information. This finding does not rule out all types of selection. For example, genetic analysis does not address our previous hypothesis that selection on stochastic epigenetic variation contributes to association between prenatal famine and BMI^16^.

Our findings were consistent across different comparison groups of famine-survivors and controls, including those born immediately before and after the famine (“time controls”) and unexposed siblings of famine survivors (“sibling controls”). Findings were also consistent using PGIs computed using the LDPred2 method,^31^ which takes account of the covariance structure of common genetic variants studied in GWAS, and a “clumping + thresholding” (C+P) approach implemented using the PRSice2 software.^29,44^

Although our results did not support substantial confounding of in-utero famine effects on obesity risk, they do motivate further consideration of gene-environment interplay. In our analysis, BMI PGIs were somewhat higher in famine survivors as compared with controls and their associations with BMI were stronger in famine exposed vs. control participants. The relatively small size of DHWFS limits statistical power to identify small rGE and GxE effects. However, larger cohorts formed by pooling DHWFS with other Dutch Famine cohorts^45^ or through genotyping of larger cohorts including substantial numbers of survivors of in-utero famine, e.g. CHARLS,^46^ could better elucidate small effects. In turn, improved understanding of small-effect G-E interplay could inform theories of how famine affects human populations as well as mechanisms through which famine influences risk for disease.

We acknowledge limitations. DHWFS was not designed to represent the full population exposed to the famine ^20^. The famine-exposed and control groups were identified from a contiguous birth series at three hospitals and were designed to form a homogenous sample that differed only with respect to famine exposure, with the goal of maximizing internal validity. However, the sample has similar characteristics to the overall population of the southern Netherlands and has produced similar estimates of famine effects when compared to population data.^47^ A related possibility is that mortality during young adulthood or midlife related to in-utero famine exposure could induce bias in the sample recruited into DHWFS and attenuation of differences between famine-survivors and controls. However, famine-related mortality up to the age at which DHWFS participants were recruited is modest (excess risk <=6%, depending on gestational timing of exposure).^48^ We observed BMI at only a single point in the life course (around age 59), while effects of famine and gene-environment interplay could vary across development. However, midlife is usually when BMI peaks and poses significant health risks, thus is a critical time to measure BMI and exam gene-environment interplay in this study. Polygenic indices are incomplete summaries of genetic risk.^43^ We used the best available methods identified in a prior, independent large-scale study.^19^ In DHWFS, this PGI explained 17% of variance in BMI, similar to published estimates from other European-descent cohorts.^49^ However, it is unknown if the genetic factors predisposing to obesity in modern cohorts had similar effects on obesity risk in the generation that composed DHWFS participants’ parents. Moreover, published estimates suggest an upper-bound to BMI PGI prediction of 30-40%^50,51^; measurement error in the PGI could therefore bias our result toward the null.

In summary, we used genetic data from a famine-natural-experiment birth cohort to evaluate potential confounding of in-utero famine effects on midlife obesity due to selective fertility and fetal survival. Results provide evidence against the presence of substantial confounding of effect estimates in genetics-naïve designs and support the internal validity of well-designed famine-natural-experiment birth-cohort studies.

By incorporating genetic analysis to evaluate selection bias, our study illustrates the potential for using genetic data to increase the robustness of observational epidemiology. Overall, this study contributes to the growing body of research investigating the effects of in-utero undernutrition on human health and identified a future direction where research with a larger cohort can continue to explore the potential small-scale effects of in-utero selection on the outcomes of natural experiments.

## Supporting information

This is a composite file for all supplemental data

## Ethics approval

This study received ethical approval from the Institutional Review Board of the Columbia University Mailman School of Public Health under protocol AAAT5074.

## Data availability

The data underlying this article are not publicly available due to privacy considerations. They can be requested through contact with the corresponding author and shared conditional on an approved research plan and proof of IRB oversight.

## Supplementary data

Supplementary data are available at IJE online.

## Author contribution

D.W.B., L.H.L., B.H., and D.C. designed and planned the study. D.W.B., L.H.L., and B.H. generated the data. J.Z., T.W., S.W., and D.W.B. analyzed the data. J.Z. and D.W.B wrote the first draft of the manuscript. All authors were involved in the interpretation of results, manuscript revision, and approval of the final version.

