## Supplementary material for "Genetic analysis of selection bias in a natural experiment: Investigating in-utero famine effects on elevated body mass index in the Dutch Hunger Winter Families Study": This is a composite file for all supplemental data

### Supplemental Method

*Polygenic Index*

To summarize the polygenic index (PGI) computation process, SNPs in the selected Genome-wide association studies (GWAS) results, Dutch Hunger Winter Families Study (DHWFS) genotype data, and HapMap3+ variants reference were first matched and extracted by taking the intersection of the three variant datasets. Next, correlation matrixes indicating the linkage equilibrium (LD) between variants were created using the HapMap3 reference data. Finally, the 'auto' model within LDPred2 requires the tuning parameters of SNP heritability ($h^2$SNP) and the prior proportion of variants assumed to be causal (p). We estimated $h^2$SNP within the LDPred2 software from LD-score regression applied to the GWAS summary statistics for each phenotype. We set the prior value of p as a sequence with length of 30 evenly spaced on a logarithmic scale from $1x10^{-4}$ to 0.3. As a sensitivity analysis, we computed a parallel set of polygenic index using a simple additive model (i.e. one with no adjustment of GWAS coefficients for patterns of LD) according to the clumping parameters determined to be optimal in the analysis by Khera et al.[1] We computed these alternative polygenic index using the PRSice2 software.[2]

*Sensitivity Analyses*

We conducted sensitivity analyses to evaluate the potential impact on results of the duration and gestational timing of famine exposure, of sex differences, of alternative methods of polygenic index construction, and to evaluate the impact of measurement error in the polygenic score. Because of the large number of tests across specifications, we adopt a p-value threshold of 0.005 to identify nominally statistically significant results following recommendations.[3]

First, we repeated our analysis focusing on the subset of famine-exposed participants who were exposed to famine during at least 10 weeks of gestation. Results were similar, although effects of famine were somewhat larger (**Table S5**).

Second, we explored the possibility that gene-environment interplay between in-utero famine exposure and genetic risk might be specific to certain periods of gestation. For this analysis, we estimated main effects of famine as well as rGE and GxE for famine exposure during each of six periods of gestation. There was no evidence of gestational-timing-specific effects (**Table S8-S9, Figure S1-S2**).

Third, we explored potential sex differences in the effects of in-utero famine. We repeated analysis separately for men and women. Results were similar for men and women (**Table S6**). Finally, we repeated analyses using an alternative specification of the BMI PGI computed using the PRSice software. Again, results were similar to those obtained using the LDpred PGI (**Tables S11-S17, Figure S3-S4**).

We conducted Simulation-Extrapolation (SIMEX) analysis to account for the measurement error in the BMI PGI. By adjusting for the measurement error in PGI, the effects were all stronger compared to effect sizes obtained from the naïve primary analyses, while did not change the conclusion of no genetic difference between famine-exposed and controls (**Table S10**).

*Sex difference sensitivity analysis*

We explored potential sex differences in findings by repeating analyses in samples stratified by sex. We conducted formal tests of sex differences by including product terms in regression models.

We explored potential heterogeneity by timing of famine exposure during gestation. We considered six windows of exposure developed based on exposure definitions in previous DHWFS studies.[4–7] For analysis of main effects of famine and gene-environment cortrealation (rGE), we replaced the single indicator of famine exposure with indicator variables identifying exposure within each of the time windows. For analysis of gene-environment interaction (GxE), we replaced the single indicator of famine exposure and the single product term with indicator variables for each exposure window and product terms for each exposure window.

*Adjustment for measurement error*

We employed the Simulation-Extrapolation (SIMEX) technique to account for the noise in the body mass index polygenic indices potentially caused by inadequate samples in the original BMI GWAS.[8,9] All effects became stronger in the SIMEX adjusted analyses compared to the results obtained from the naïve primary models (**Table S10**). However, the SIMEX adjusted results still support our findings of no large-scale selection was in place to bias the effect of famine on midlife BMI in the DHWFS cohort. The intensification of SIMEX adjusted effect sizes suggests that our power to rule out selection bias in the DHWFS cohort could be limited and reproduce our analyses in larger sample with increased statistical power should be considered in the future. SIMEX analyses were conducted with R (version 4.1.2) using the `simex` (version 1.8) package.[10,11]

**Table S1. Sample Characteristics.** The table shows the characteristics of the analysis sample (left side) and the full Dutch Hunger Winter Families Study (DHWFS) Cohort (right side). The data in the table show that our analysis sample is comparable to the full DHWFS sample.

| | DHWFS Cohort (N = 1031) | | | | Analysis Sample (N = 950) | | | |
|---|---|---|---|---|---|---|---|---|
| | N | (%) | Mean | SD[1] | N | (%) | Mean | (SD) |
| **Exposure Group** | | | | | | | | |
| Exposed | 557 | (53%) | | | 485 | (51%) | | |
| Time controls | 176 | (17%) | | | 161 | (16%) | | |
| Sib controls | 308 | (30%) | | | 204 | (32%) | | |
| **Age at Telephone Interview** | 1,031 | | 59 | (4) | 950 | | 59 | (4) |
| **Gender** | | | | | | | | |
| Men | 465 | (45%) | | | 430 | (45%) | | |
| Women | 566 | (54%) | | | 520 | (54%) | | |
| **Health Outcome (measured in the sample participating in the clinic exam)** | | | | | | | | |
| BMI (n=968) | 968 | | 27.7 | (4.6) | 947 | | 27.7 | (4.6) |

[1] SD: Standard Deviation

**Table S2. Definition of six timing-specific famine exposure groups with their cutoff dates based on the last menstrual period.**

| Names | Exposed groups | Start date | End date |
|---|---|---|---|
| D4 | Exposed in weeks 31 to delivery | Apr 20, 1944[#] | Aug 24, 1944 |
| D3 | Exposed in weeks 21 to 30 | Jul 9, 1944 | Oct 15, 1944 |
| D2 | Exposed in weeks 11 to 20 | Sept 17, 1944 | Dec 24, 1944 |
| D1 | Exposed in weeks 1 to 10 | Nov 26, 1944 | Mar 4, 1945 |
| D0 | Exposed in weeks -9 to 0 (conception to less than 10 weeks) | Feb 4, 1945 | May 12, 1945 |
| D-1 | Exposed in preconceptional exposure | May 13, 1945 | June 25, 1945 |

*The start and end dates included for each period.
#Updated start date was calculated as Apr 30, 1944; however, we added 9 postnatal exposed individuals to D4, which makes the start date Apr 20, 1944

**Supplemental Results produced with LDpred2 Polygenic Indices:**

**Table S3. LDpred2 Polygenic Index (PGI) Summary Statistics.** The table shows the mean and 95%CI for BMI PGI computed based on three selected genome-wide association study results (GWAS), for the analysis sample. The analysis sample was consisted of three groups: the famine-exposed group, the time-controls sampled from births at the same hospitals before and after the famine period, and unexposed same-sex siblings of the famine-exposed and time-control participants. This PGI were computed using LDpred2 software with the 'auto' model.

*Summary Statistics of Polygenic Index (N = 950, LDpred2 indices)*

| | Famine-exposed (N = 485) | | Time Controls (N = 161) | | Sibling Controls (N = 304) | |
|---|---|---|---|---|---|---|
| | Mean | 95%CI[1] | Mean | 95%CI | Mean | 95%CI |
| GWAS: | | | | | | |
| Body Mass Index (BMI) | 0.04 | (-0.69, 0.79) | 0.00 | (-0.62, 0.69) | -0.07 | (-0.72, 0.52) |

[1] CI: Confidence Interval

**Table S4. Genetic analysis of in-utero famine exposure effects on Body Mass Index (BMI).** The table shows results from regression analysis of BMI. **Panel A** reports analysis of main effects of famine exposure and genetic risk on BMI. Effect-sizes for famine exposure and genetic risk were estimated in separate models. The “Primary Analysis” model included all participants in our analysis sample with available data on BMI (n=947) and was fitted using linear regression within a generalized estimating equations (GEE) framework to account for non-independence of data from sibling pairs. Models included covariates for participant age, age-squared, and sex. The “Sibling Difference” model included the subset of the Primary Analysis sample consisting of sibling pairs discordant for famine exposure (n=226 pairs) and was fitted using fixed effects regression. Models included covariates for age and age-squared (all sibling pairs were of the same sex). The “Exposed vs. Time Control” model included the subset of the Primary Analysis sample consisting of famine exposure participants and their time controls (n= 644) and was fitted using linear regression within a generalized estimating equations (GEE) framework. Effect-sizes are reported as regression beta coefficient and 95% confidence intervals (CI). Genetic association between BMI PGI and BMI were also reported in Panel A. Similar to models assessing main effect of genetic risk which regressed BMI PGI onto BMI, genetic association between BMI PGI and BMI was assessed in three samples: “Famine-exposed” (n = 485), “Time Controls” (n = 161), “Sibling Controls” of famine-exposed (simplified as “Sibling Controls” in the plot legend: n = 226), and “All Controls” consist of time controls, sibling controls of time controls, and sibling controls of famine-exposed (n = 465).

**Panel B** reports analysis of gene-environment correlation (rGE) for BMI. The “Primary Analysis” model included all participants in our analysis sample (n=950) and was fitted using linear regression within a generalized estimating equations (GEE) framework to account for non-independence of data from sibling pairs. Models included covariates for participant age, age-squared, and sex. The “Sibling Difference” model included the subset of the Primary Analysis sample consisting of sibling pairs discordant for famine exposure (n=226 pairs) and was fitted using linear fixed effects regression. Models included covariates for age and age-squared (all sibling pairs were of the same sex). The “Exposed vs. Time Control” model included the subset of the Primary Analysis sample consisting of famine exposure participants and their time controls (n= 646) and was fitted using linear regression within a generalized estimating equations (GEE) framework. All rGE effect-sizes were a regression beta coefficient interpretable as Cohen’s d (because the PGI is analyzed on standardized scale). **Panel C** reports analysis of gene-environment interaction (GxE) for BMI. Similar as analyses in panel A, a “Primary Analysis” model (n = 934), a “Sibling Difference” model (n=220 pairs), and a “Exposed vs. Time Control” model (n = 632) were constructed with different sub-samples selected from the analysis sample. In each model, coefficients are reported for model terms for famine exposure, genetic risk, and a product term testing their interaction. Coefficients are reported as beta coefficient estimated from linear regression fitted using GEE framework to account for non-independence of sibling data. Models included covariates for age, age-squared, and sex.

| Body Mass Index | | | |
|---|---|---|---|
| **Panel A** | | | |
| | Beta | 95% CI | p-value |
| Main Effect of Famine | | | |
| Primary Analysis | 0.28 | 0.16, 0.40 | <0.001 |
| Exposed vs. Time Control | 0.25 | 0.06, 0.43 | 0.009 |
| Sibling Difference | 0.29 | 0.12, 0.46 | <0.001 |
| Main Effect of Famine (adjusted by PGI) | | | |
| Primary Analysis | 0.23 | 0.12, 0.34 | <0.001 |
| Exposed vs. Time Control | 0.23 | 0.06, 0.40 | 0.008 |
| Sibling Difference | 0.27 | 0.12, 0.43 | <0.001 |
| Main Effect of PGI | | | |
| Primary Analysis | 0.42 | 0.35, 0.49 | <0.001 |
| Exposed vs. Time Control | 0.41 | 0.32, 0.49 | <0.001 |
| Sibling Difference | 0.46 | 0.31, 0.61 | <0.001 |
| Genetic Association with BMI | | | |
| Famine-exposed | 0.43 | 0.34, 0.52 | <0.001 |
| Time Controls | 0.33 | 0.19, 0.46 | <0.001 |
| Sibling Controls | 0.39 | 0.28, 0.50 | <0.001 |
| All Controls | 0.38 | 0.30, 0.47 | <0.001 |
| **Panel B** | | | |
| | Beta | 95% CI | p-value |
| rGE | | | |
| Primary Analysis | 0.10 | -0.02, 0.21 | 0.092 |
| Exposed vs. Time Control | 0.04 | -0.14, 0.22 | 0.674 |
| Sibling Difference | 0.04 | -0.11, 0.18 | 0.601 |
| **Panel C:** | | | |
| | Beta | 95% CI | p-value |
| GxE | | | |
| Primary Analysis | | | |
| Famine | 0.23 | 0.12, 0.34 | <0.001 |
| PGI | 0.39 | 0.30, 0.47 | <0.001 |
| Famine* PGI | 0.05 | -0.08, 0.17 | 0.457 |
| Exposed vs. Time Control | | | |
| Famine | 0.23 | 0.06, 0.39 | 0.009 |
| PGI | 0.34 | 0.19, 0.49 | <0.001 |
| Famine* PGI | 0.10 | -0.08, 0.27 | 0.272 |
| Sibling Difference | | | |

| | | | |
|---|---|---|---|
| Famine | 0.28 | 0.12, 0.44 | <0.001 |
| PGI | 0.41 | 0.26, 0.57 | <0.001 |
| Famine* PGI | 0.07 | -0.08, 0.23 | 0.363 |

**Table S5. Genetic analysis of in-utero famine exposure effects on Body Mass Index (BMI) adjusted for less than 10 weeks and preconceptual famine exposure (D-1 and D0).** The table shows results from regression analysis of BMI adjusted by exposure to D-1 and D0 (details see Supplemental Table 2). **Panel A** reports analysis of main effects of famine exposure and genetic risk on BMI. Effect-sizes for famine exposure adjusted by D-1 and D0, and genetic risk were estimated in separate models. The “Primary Analysis” model included all participants in our analysis sample with available data on BMI (n=947) and was fitted using linear regression within a generalized estimating equations (GEE) framework to account for non-independence of data from sibling pairs. Models included covariates for D-1, D0, participant age, age-squared, and sex. The “Sibling Difference” model included the subset of the Primary Analysis sample consisting of sibling pairs discordant for famine exposure (n=226 pairs) and was fitted using fixed effects regression. Models included covariates for D-1, D0, age and age-squared (all sibling pairs were of the same sex). The “Exposed vs. Time Control” model included the subset of the Primary Analysis sample consisting of famine exposure participants and their time controls (n= 644) and was fitted using linear regression within a generalized estimating equations (GEE) framework. Effect-sizes are reported as regression beta coefficient and 95% confidence intervals (CI). Genetic association between BMI PGI and BMI were also reported in Panel A. Similar to models assessing main effect of genetic risk which regressed BMI PGI onto BMI, genetic association between BMI PGI and BMI was assessed in three samples: “Famine-exposed” (n = 485), “Time Controls” (n = 161), “Sibling Controls” of famine-exposed (simplified as “Sibling Controls” in the plot legend: n = 226), and “All Controls” consist of time controls, sibling controls of time controls, and sibling controls of famine-exposed (n = 465).

**Panel B** reports analysis of gene-environment correlation (rGE) for BMI. The “Primary Analysis” model included all participants in our analysis sample (n=950) and was fitted using linear regression within a generalized estimating equations (GEE) framework to account for non-independence of data from sibling pairs. Models included covariates for D-1, D0, participant age, age-squared, and sex. The “Sibling Difference” model included the subset of the Primary Analysis sample consisting of sibling pairs discordant for famine exposure (n=226 pairs) and was fitted using linear fixed effects regression. Models included covariates for D-1, D0, age and age-squared (all sibling pairs were of the same sex). The “Exposed vs. Time Control” model included the subset of the Primary Analysis sample consisting of famine exposure participants and their time controls (n= 646) and was fitted using linear regression within a generalized estimating equations (GEE) framework. All rGE effect-sizes were a regression beta coefficient interpretable as Cohen’s d (because the PGI is analyzed on standardized scale). **Panel C** reports analysis of gene-environment interaction (GxE) for BMI. Similar as analyses in panel A, a “Primary Analysis” model (n = 934), a “Sibling Difference” model (n=220 pairs), and a “Exposed vs. Time Control” model (n = 632) were constructed with different sub-samples selected from the analysis sample. In each model, coefficients are reported for model terms for famine exposure, genetic risk, and a product term testing their interaction. Coefficients are reported as beta coefficient estimated from linear regression fitted using GEE framework to account for non-independence of sibling data. Models included covariates for D-1, D0, age, age-squared, and sex.

| Body Mass Index | | | |
|---|---|---|---|
| **Panel A** | | | |
| | Beta | 95% CI | p-value |
| Main Effect of Famine | | | |
| Primary Analysis | 0.32 | 0.19, 0.46 | <0.001 |
| Exposed vs. Time Control | 0.30 | 0.10, 0.49 | 0.003 |
| Sibling Difference | 0.33 | 0.14, 0.52 | <0.001 |
| Main Effect of Famine (adjusted by PGI) | | | |
| Primary Analysis | 0.28 | 0.15, 0.40 | <0.001 |
| Exposed vs. Time Control | 0.28 | 0.10, 0.46 | 0.002 |
| Sibling Difference | 0.31 | 0.13, 0.49 | <0.001 |
| Main Effect of PGI | | | |
| Primary Analysis | 0.42 | 0.35, 0.49 | <0.001 |
| Exposed vs. Time Control | 0.41 | 0.34, 0.49 | <0.001 |
| Sibling Difference | 0.46 | 0.31, 0.60 | <0.001 |
| Genetic Association with BMI | | | |
| Famine-exposed | 0.43 | 0.34, 0.52 | <0.001 |
| Time Controls | 0.33 | 0.19, 0.46 | <0.001 |
| Sibling Controls | 0.39 | 0.28, 0.50 | <0.001 |
| All Controls | 0.38 | 0.30, 0.47 | <0.001 |
| **Panel B** | | | |
| | Beta | 95% CI | p-value |
| rGE | | | |
| Primary Analysis | 0.08 | -0.04, 0.21 | 0.174 |
| Exposed vs. Time Control | 0.03 | -0.16, 0.21 | 0.781 |
| Sibling Difference | 0.05 | -0.11, 0.22 | 0.516 |
| **Panel C:** | | | |
| | Beta | 95% CI | p-value |
| GxE | | | |
| Primary Analysis | | | |
| Famine | 0.28 | 0.16, 0.40 | <0.001 |
| PGI | 0.39 | 0.30, 0.47 | <0.001 |
| Famine* PGI | 0.04 | -0.10, 0.19 | 0.541 |
| Exposed vs. Time Control | | | |
| Famine | 0.28 | 0.10, 0.46 | 0.002 |
| PGI | 0.34 | 0.19, 0.49 | <0.001 |
| Famine* PGI | 0.09 | -0.09, 0.27 | 0.341 |
| Sibling Difference | | | |

| | | | |
|---|---|---|---|
| Famine | 0.32 | 0.14, 0.49 | <0.001 |
| PGI | 0.43 | 0.27, 0.58 | <0.001 |
| Famine* PGI | 0.17 | -0.01, 0.35 | 0.071 |

**Table S6. Sex stratified genetic analysis, with LDpred2 generated polygenetic index, of in-utero famine exposure effects on Body Mass Index (BMI). Panel A** reports analysis of main effects of famine exposure and genetic risk on BMI. Effect-sizes for famine exposure and genetic risk were estimated in separate models. The "Primary Analysis" model included all participants in our analysis sample with available data on BMI (n=947) and was fitted using linear regression within a generalized estimating equations (GEE) framework to account for non-independence of data from sibling pairs. Models included covariates for participant age, age-squared, and sex. We repeated the main effect analysis in the "Women" (n = 520) and "Men" (n = 427 ) strata separately. Models included covariates for age and age squared. Tests of sex difference were conducted . Coefficients are reported for two product terms: famine and sex, and PGI and sex. Sex interaction coefficients are reported as beta estimates from linear regression fitted using GEE framework to account for non-independence of sibling data. **Panel B** reports analysis of gene-environment correlation (rGE) for BMI polygenic indices. The effect-size is estimated from a linear regression of the BMI PGI on famine exposure fitted using GEE to account for non-independence of data from siblings and including covariates for age, age-squared, and sex. The effect-size is a regression beta coefficient interpretable as Cohen's d (because the PGI is analyzed on standardized scale). We repeated the rGE analysis among women and men separately. Models included covariates for age and age squared. Test of sex difference was conducted. Coefficients are reported for models' interaction term between famine and sex. Sex interaction coefficient is reported as beta estimates from linear regression fitted using GEE framework to account for non-independence of sibling data. **Panel C** reports analysis of gene-environment interaction (GxE) for BMI. Coefficients are reported for model terms for famine exposure, genetic risk, and the product term testing super-multiplicative interaction. Coefficients are reported as beta estimated from linear regression fitted using GEE framework to account for non-independence of sibling data. Models included covariates for age, age-squared, and sex. We repeated the GxE analysis among women and man separately. Models included covariates for age and age squared. Test of sex difference was conducted. Coefficients are reported for three model product terms testing super-multiplicative interaction between famine, PGI, and sex. These interaction coefficients are reported as beta estimates from linear regression fitted using GEE framework to account for non-independence of sibling data.

| | Body Mass Index | | |
|---|---|---|---|
| **Panel A:** | | | |
| | Beta | 95% CI | p-values |
| Main Effect of Famine | | | |
| Primary Analysis | 0.28 | 0.16, 0.40 | <0.001 |
| Women | 0.39 | 0.21, 0.57 | <0.001 |
| Men | 0.14 | -0.01, 0.30 | 0.070 |
| Test of Sex Difference | -0.20 | -0.42, 0.02 | 0.069 |
| Main Effect of PGI | | | |
| Primary Analysis | 0.42 | 0.35, 0.49 | <0.001 |
| Women | 0.48 | 0.38, 0.58 | <0.001 |
| Men | 0.33 | 0.24, 0.43 | <0.001 |
| Test of Sex Difference | -0.15 | -0.28, -0.02 | 0.029 |
| **Panel B:** | | | |
| | Beta | 95% CI | p-value |
| rGE | | | |
| Primary Analysis | 0.10 | -0.02, 0.21 | 0.092 |

| | | | |
|---|---|---|---|
| Women | 0.17 | 0.02, 0.32 | 0.029 |
| Men | 0.01 | -0.17, 0.17 | 0.959 |
| Test of Sex Difference | -0.20 | -0.41, 0.01 | 0.057 |

**Panel C:**

| | Beta | 95%CI | p-value |
|---|---|---|---|
| **GxE** | | | |
| Primary Analysis | | | |
| Famine | 0.23 | 0.12, 0.34 | <0.001 |
| PGI | 0.39 | 0.30, 0.47 | <0.001 |
| Famine: PGI | 0.05 | -0.08, 0.17 | 0.457 |
| Women | | | |
| Famine | 0.30 | 0.14, 0.46 | <0.001 |
| PGI | 0.44 | 0.31, 0.56 | <0.001 |
| Famine: PGI | 0.06 | -0.12, 0.23 | 0.510 |
| Men | | | |
| Famine | 0.14 | -0.01, 0.28 | 0.069 |
| PGI | 0.32 | 0.22, 0.42 | <0.001 |
| Famine: PGI | 0.03 | -0.14, 0.20 | 0.757 |
| Test of Sex Difference | | | |
| Famine:Sex | -0.11 | -0.31, 0.10 | 0.304 |
| PGI:Sex | -0.12 | -0.28, 0.04 | 0.131 |
| Famine: PGI:Sex | -0.02 | -0.27, 0.22 | 0.842 |

**Table S7. Sex stratified genetic analysis adjusted by preconceptual exposure groups (D-1 and D0), with LDpred2 generated polygenetic index, of in-utero famine exposure effects on Body Mass Index (BMI). Panel A** reports analysis of main effects of famine exposure and genetic risk on BMI. Effect-sizes for famine exposure and genetic risk were estimated in separate models. The "Primary Analysis" model included all participants in our analysis sample with available data on BMI (n=947) and was fitted using linear regression within a generalized estimating equations (GEE) framework to account for non-independence of data from sibling pairs. Models included covariates for D-1, D0, participant age, age-squared, and sex. We repeated the main effect analysis in the "Women" (n = 520) and "Men" (n = 427 ) strata separately. Models included covariates for D-1, D0, age and age squared. Tests of sex difference were conducted . Coefficients are reported for two product terms: famine and sex, and PGI and sex. Sex interaction coefficients are reported as beta estimates from linear regression fitted using GEE framework to account for non-independence of sibling data. **Panel B** reports analysis of gene-environment correlation (rGE) for BMI polygenic indices. The effect-size is estimated from a linear regression of the BMI PGI on famine exposure fitted using GEE to account for non-independence of data from siblings and including covariates for D-1, D0, age, age-squared, and sex. The effect-size is a regression beta coefficient interpretable as Cohen's d (because the PGI is analyzed on standardized scale). We repeated the rGE analysis among women and men separately. Models included covariates for D-1, D0, age and age squared. Test of sex difference was conducted. Coefficients are reported for models' interaction term between famine and sex. Sex interaction coefficient is reported as beta estimates from linear regression fitted using GEE framework to account for non-independence of sibling data. **Panel C** reports analysis of gene-environment interaction (GxE) for BMI. Coefficients are reported for model terms for famine exposure, genetic risk, and the product term testing super-multiplicative interaction. Coefficients are reported as beta estimated from linear regression fitted using GEE framework to account for non-independence of sibling data. Models included covariates for D-1, D0, age, age-squared, and sex. We repeated the GxE analysis among women and man separately. Models included covariates for age and age squared. Test of sex difference was conducted. Coefficients are reported for three model product terms testing super-multiplicative interaction between famine, PGI, and sex. These interaction coefficients are reported as beta estimates from linear regression fitted using GEE framework to account for non-independence of sibling data.

| Body Mass Index | | | |
|---|---|---|---|
| **Panel A:** | | | |
| | Beta | 95% CI | p-values |
| Main Effect of Famine | | | |
| Primary Analysis | 0.32 | 0.19, 0.46 | <0.001 |
| Women | 0.50 | 0.30, 0.70 | <0.001 |
| Men | 0.12 | -0.05, 0.28 | 0.172 |
| Test of Sex Difference | -0.20 | -0.42, 0.02 | 0.069 |
| Main Effect of PGI | | | |
| Primary Analysis | 0.42 | 0.35, 0.49 | <0.001 |
| Women | 0.48 | 0.38, 0.58 | <0.001 |
| Men | 0.33 | 0.24, 0.43 | <0.001 |
| Test of Sex Difference | -0.15 | -0.28, -0.02 | 0.029 |
| **Panel B:** | | | |
| | Beta | 95% CI | p-value |
| rGE | | | |

| | Beta | 95%CI | p-value |
|---|---|---|---|
| Primary Analysis | 0.08 | -0.04, 0.21 | 0.174 |
| Women | 0.15 | -0.01, 0.31 | 0.073 |
| Men | -0.01 | -0.19, 0.17 | 0.895 |
| Test of Sex Difference | -0.20 | -0.41, 0.01 | 0.057 |

**Panel C:**

| | Beta | 95%CI | p-value |
|---|---|---|---|
| **GxE** | | | |
| Primary Analysis | | | |
| Famine | 0.28 | 0.16, 0.40 | <0.001 |
| PGI | 0.39 | 0.30, 0.47 | <0.001 |
| Famine: PGI | 0.04 | -0.10, 0.19 | 0.541 |
| Women | | | |
| Famine | 0.41 | 0.23, 0.59 | <0.001 |
| PGI | 0.44 | 0.31, 0.56 | <0.001 |
| Famine: PGI | 0.07 | -0.14, 0.28 | 0.498 |
| Men | | | |
| Famine | 0.12 | -0.05, 0.28 | 0.158 |
| PGI | 0.32 | 0.22, 0.42 | <0.001 |
| Famine: PGI | -0.01 | -0.20, 0.19 | 0.956 |
| Test of Sex Difference | | | |
| Famine:Sex | -0.11 | -0.31, 0.10 | 0.304 |
| PGI:Sex | -0.12 | -0.28, 0.04 | 0.131 |
| Famine: PGI:Sex | -0.02 | -0.27, 0.22 | 0.842 |

**Table S8. Sensitivity analysis on the timing-specific gene-environment correlation between in-utero famine exposure and LDpred2 produced body mass index (BMI) polygenic index (PGI).** This table report the analysis of timing-specific gene-environment correlation between famine exposure and genetic risk on BMI. Effect-sizes for six famine exposure groups (D4 ~ D-1) were estimated in a model which included all participants in our analysis sample with available data on BMI (n=950) and was fitted using linear regression within a generalized estimating equations (GEE) framework to account for non-independence of data from sibling pairs. Model regressed genetic risk of elevated BMI onto six famine exposure groups and adjusted for covariates of participant age, age-squared, and sex.

| | Timing-specific gene-environment correlation | | |
|---|---|---|---|
| | Beta | 95% CI | p-values |
| D4 (weeks 31 to delivery) | -0.02 | -0.20, 0.16 | 0.848 |
| D3 (gestational weeks 21-30) | 0.14 | -0.03, 0.31 | 0.101 |
| D2 (gestational weeks 11-20) | -0.02 | -0.20, 0.15 | 0.794 |
| D1 (gestational weeks 1-10) | 0.18 | -0.04, 0.41 | 0.111 |
| D0 (conception to less than 10 weeks) | 0.04 | -0.18, 0.27 | 0.699 |
| D-1 (preconceptional) | 0.18 | -0.06, 0.42 | 0.147 |

**Figure S1. Sensitivity analysis on the timing-specific gene-environment correlation between in-utero famine exposure and LDpred2 produced body mass index (BMI) polygenic index (PGI).** This plot shows the correlation between genetic risk and timing-specific famine exposures (D4 ~ D-1). Timing of famine exposure was shown on the x-axis. The beta effect sizes estimated from linear regression of the BMI PGI on famine exposure with six time-specific exposure windows added as covariates were shown on the y-axis . Model was fitted using generalized estimating equations (GEE) framework to account for non-independence of data from sibling pairs.

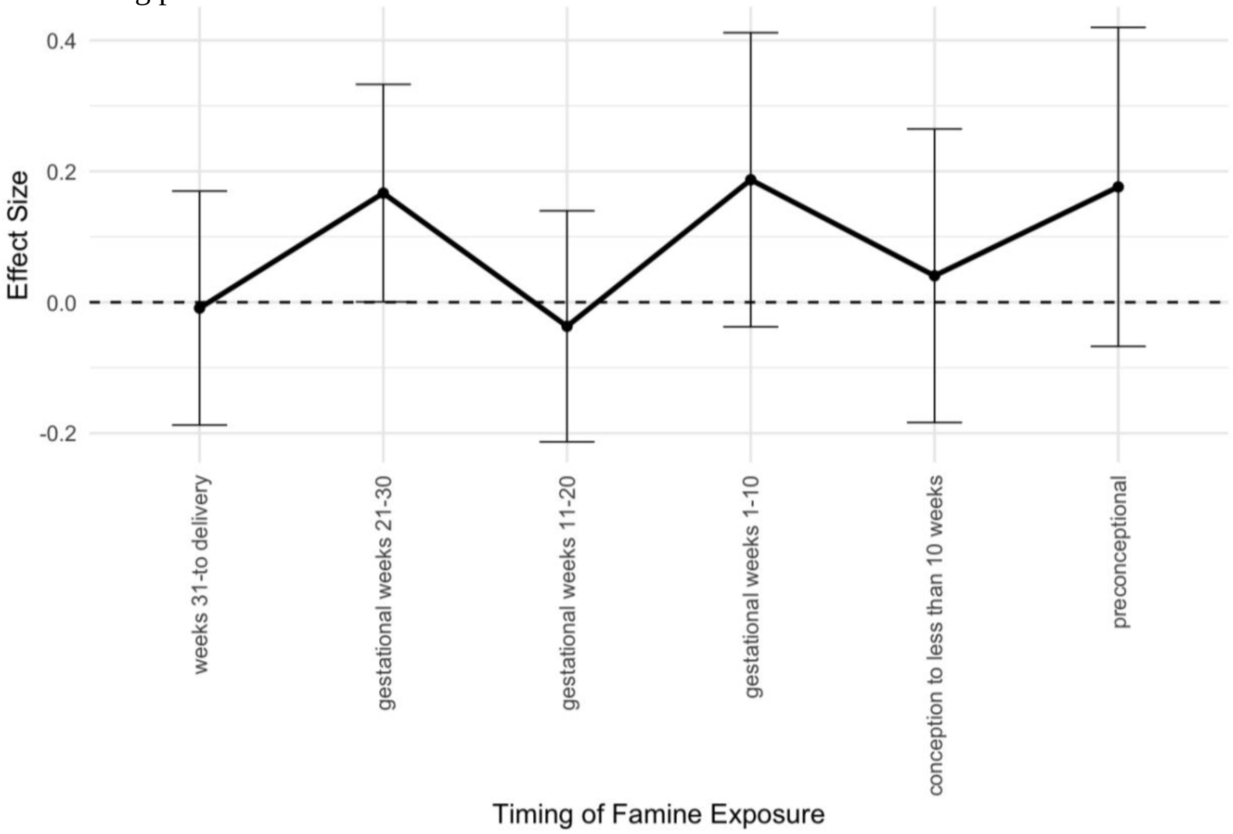

**Table S9. Sensitivity analysis on the timing-specific gene-environment interaction between in-utero famine exposure and LDpred2 produced body mass index (BMI) polygenic index (PGI).** This table report the analysis of timing-specific gene-environment interaction between famine exposure and genetic risk on BMI. Effect-sizes for six product terms between famine exposure groups (D4 ~ D-1)and genetic risk of elevated BMI were estimated in a model which included all participants in our analysis sample with available data on BMI (n=950) and was fitted using linear regression within a generalized estimating equations (GEE) framework to account for non-independence of data from sibling pairs. Model regressed BMI onto six product terms between famine exposure groups and genetic risk of elevated BMI. Model was adjusted for covariates of participant age, age-squared, and sex.

| Timing-specific gene-environment interaction | | | |
|---|---|---|---|
| | Beta | 95% CI | p-values |
| PGI:D4 (weeks 31 to delivery) | 0.27 | 0.07, 0.47 | 0.008 |
| PGI:D3 (gestational weeks 21-30) | 0.25 | 0.04, 0.45 | 0.019 |
| PGI:D2 (gestational weeks 11-20) | 0.30 | 0.07, 0.53 | 0.012 |
| PGI:D1 (gestational weeks 1-10) | 0.27 | 0.02, 0.52 | 0.032 |
| PGI:D0 (conception to less than 10 weeks) | 0.43 | 0.27, 0.58 | <0.001 |
| PGI:D-1 (preconceptional) | 0.26 | -0.11, 0.62 | 0.167 |

**Table S10. Regression estimates from primary analyses and SIMEX adjusted estimates.** The "Primary Analysis" models included all participants in our analysis sample with available data on BMI (n=947) and was fitted using linear regression within a generalized estimating equations (GEE) framework to account for non-independence of data from sibling pairs. All models included covariates for participant age, age-squared, and sex. The first model reports the main effects of genetic risk on BMI. The gene-environment correlation (rGE) reports the association between BMI PGI and famine exposure status. For the gene-environment interaction (GxE) regression model, the coefficients are reported for model terms for famine exposure, genetic risk, and a product term testing their interaction. Effect-sizes are reported as regression beta coefficient and standard deviation in the parenthesis. The "SIMEX Adjusted Analysis" models were constructed based on the naive models in the "Primary Analyses" with an additional measurement error adjustment term. SIMEX-adjusted effect-sizes are reported as regression beta coefficient and standard deviation in the parenthesis.

| | Primary Analysis | SIMEX Adjusted Analyses |
|---|---|---|
| Main Effect of PGI | 0.42***<br>(0.035) | 0.51***<br>(0.042) |
| rGE | 0.10<br>(0.058) | 0.13<br>(0.083) |
| GxE | | |
| Famine | 0.23***<br>(0.056) | 0.20**<br>(0.062) |
| PGI | 0.39***<br>(0.044) | 0.47***<br>(0.062) |
| Famine:PGI | 0.047<br>(0.063) | 0.06<br>(0.090) |
| Standard errors in parentheses.<br>*** p< .001; ** p < .01; * p < .05 | | |

**Figure S2. Sensitivity analysis on the timing-specific gene-environment interaction between in-utero famine exposure and LDpred2 produced body mass index (BMI) polygenic index (PGI).** This plot shows the interaction between genetic risk and timing-specific famine exposures. Timing of famine exposure was shown on the x-axis. The beta coefficients of interaction terms between genetic risk of elevated BMI and the six famine exposure groups (D4 ~ D-1) representing the genetic effect among the famine exposed group were shown on the y-axis. These beta coefficients were estimated from linear regression of the BMI PGI on famine exposure with six product terms between PGI and the exposure windows added as covariates. Model was fitted using generalized estimating equations (GEE) framework to account for non-independence of data from sibling pairs. The grey line is showing the reference main effect of genetic risk for BMI without adjusting for timing-specific covariates, which was obtained in the primary main genetic effect analysis.

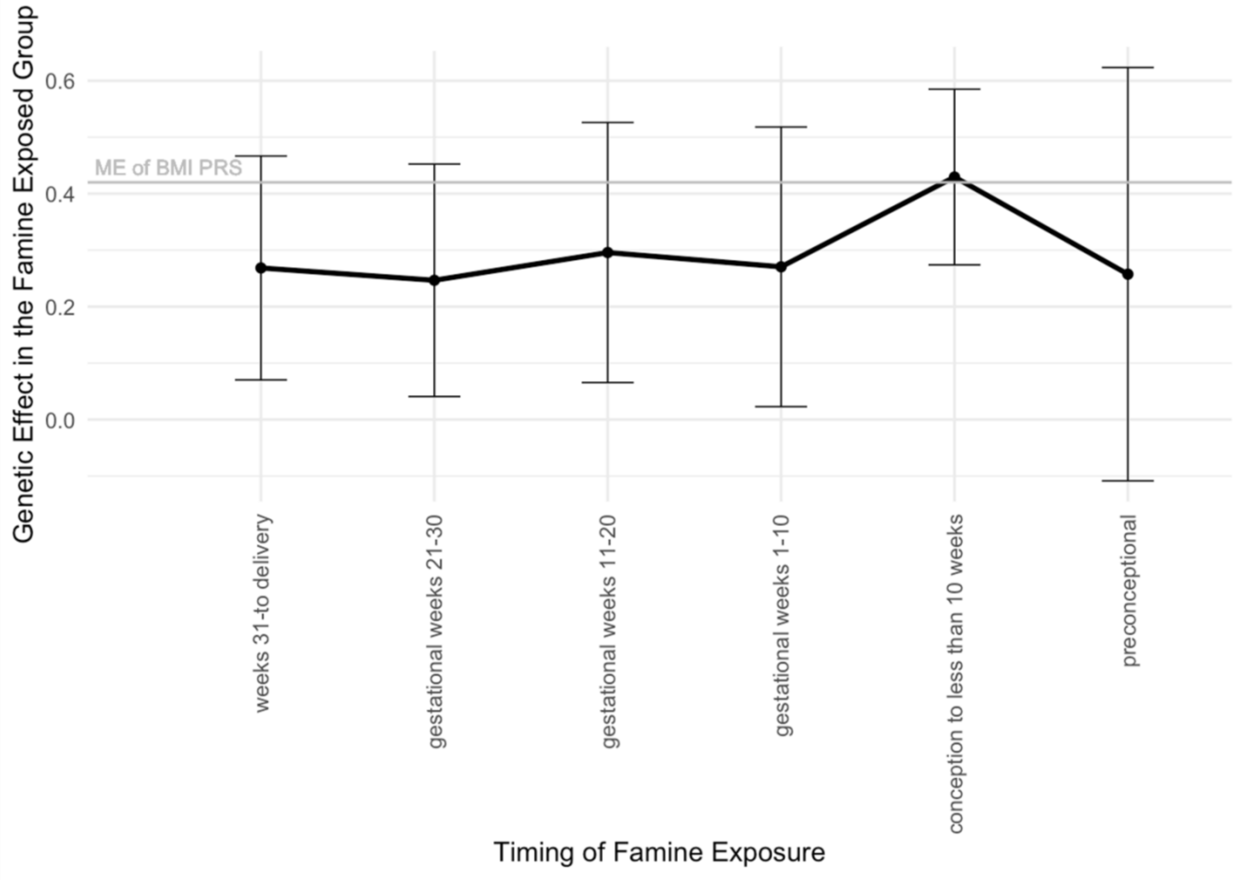

**Supplemental Results produced with PRSice2 Polygenic Indices:**

**Table S11. PRSice2 Polygenic Index (PGI) Summary Statistics.** The table shows the mean and 95%CI for the PGI computed based on selected body mass index genome-wide association study result, for the analysis sample. The analysis sample was consisted of three groups: the famine-exposed group, the time-controls sampled from births at the same hospitals before and after the famine period, and unexposed same-sex siblings of the famine-exposed and time-control participants. This PGI were computed using PRSice2 software.

*Summary Statistics of Polygenic Index (N = 950, PRSice2 indices)*

| | Famine-exposed (N = 485) | | Time Controls (N = 161) | | Sibling Controls (N = 304) | |
|---|---|---|---|---|---|---|
| | Mean | 95%CI[1] | Mean | 95%CI | Mean | 95%CI |
| GWAS: | | | | | | |
| BMI | 0.02 | (-0.65, 0.69) | 0.04 | (-0.52, 0.81) | -0.09 | (-0.74, 0.57) |

[1] CI: Confidence Interval

**Table S12. Genetic analysis with PRSice2 generated polygenetic index (PGI) of in-utero famine exposure effects on Body Mass Index (BMI).** The table shows results from regression analysis of BMI. **Panel A** reports analysis of main effects of famine exposure and genetic risk on BMI. Effect-sizes for famine exposure and genetic risk were estimated in separate models. The "Primary Analysis" model included all participants in our analysis sample with available data on BMI (n=947) and was fitted using linear regression within a generalized estimating equations (GEE) framework to account for non-independence of data from sibling pairs. Models included covariates for participant age, age-squared, and sex. The "Sibling Difference" model included the subset of the Primary Analysis sample consisting of sibling pairs discordant for famine exposure (n=226 pairs) and was fitted using fixed effects regression. Models included covariates for age and age-squared (all sibling pairs were of the same sex). The "Exposed vs. Time Control" model included the subset of the Primary Analysis sample consisting of famine exposure participants and their time controls (n= 644) and was fitted using linear regression within a generalized estimating equations (GEE) framework. Effect-sizes are reported as regression beta coefficient and 95% confidence intervals (CI). Genetic association between BMI PGI and BMI were also reported in Panel A. Similar to models assessing main effect of genetic risk which regressed BMI PGI onto BMI, genetic association between BMI PGI and BMI was assessed in three samples: "Famine-exposed" (n = 485), "Time Controls" (n = 161), "Sibling Controls" of famine-exposed (simplified as "Sibling Controls" in the plot legend: n = 226), and "All Controls" consist of time controls, sibling controls of time controls, and sibling controls of famine-exposed (n = 465). **Panel B** reports analysis of gene-environment correlation (rGE) for BMI. The "Primary Analysis" model included all participants in our analysis sample (n=950) and was fitted using linear regression within a generalized estimating equations (GEE) framework to account for non-independence of data from sibling pairs. Models included covariates for participant age, age-squared, and sex. The "Sibling Difference" model included the subset of the Primary Analysis sample consisting of sibling pairs discordant for famine exposure (n=226 pairs) and was fitted using linear fixed effects regression. Models included covariates for age and age-squared (all sibling pairs were of the same sex). The "Exposed vs. Time Control" model included the subset of the Primary Analysis sample consisting of famine exposure participants and their time controls (n= 646) and was fitted using linear regression within a generalized estimating equations (GEE) framework. All rGE effect-sizes were a regression beta coefficient interpretable as Cohen's d (because the PGI is analyzed on standardized scale). **Panel C** reports analysis of gene-environment interaction (GxE) for BMI. Similar as analyses in panel A, a "Primary Analysis" model (n = 934), a "Sibling Difference" model (n=220 pairs), and a "Exposed vs. Time Control" model (n = 632) were constructed with different sub-samples selected from the analysis sample. In each model, coefficients are reported for model terms for famine exposure, genetic risk, and a product term testing their interaction. Coefficients are reported as beta coefficient estimated from linear regression fitted using GEE framework to account for non-independence of sibling data. Models included covariates for age, age-squared, and sex.

**Body Mass Index**

**Panel A**

| | Beta | 95% CI | p-value |
|---|---|---|---|
| Main Effect of Famine | | | |
| Primary Analysis | 0.28 | 0.16, 0.40 | <0.001 |
| Exposed vs. Time Control | 0.25 | 0.06, 0.43 | 0.009 |
| Sibling Difference | 0.29 | 0.12, 0.46 | <0.001 |
| Main Effect of Famine (adjusted by PGI) | | | |
| Primary Analysis | 0.25 | 0.13, 0.37 | <0.001 |
| Exposed vs. Time Control | 0.26 | 0.09, 0.44 | 0.003 |
| Sibling Difference | 0.26 | 0.09, 0.43 | 0.002 |
| Main Effect of PGI | | | |
| Primary Analysis | 0.31 | 0.24, 0.37 | <0.001 |
| Exposed vs. Time Control | 0.31 | 0.23, 0.38 | <0.001 |
| Sibling Difference | 0.25 | 0.10, 0.40 | <0.001 |
| Genetic Association with BMI | | | |
| Famine-exposed | 0.33 | 0.24, 0.42 | <0.001 |
| Time Controls | 0.23 | 0.09, 0.37 | 0.002 |
| Sibling Controls | 0.25 | 0.13, 0.37 | <0.001 |
| All Controls | 0.28 | 0.19, 0.36 | <0.001 |

**Panel B**

| | Beta | 95% CI | p-value |
|---|---|---|---|
| rGE | | | |
| Primary Analysis | 0.09 | -0.03, 0.20 | 0.152 |
| Exposed vs. Time Control | -0.07 | -0.25, 0.11 | 0.433 |
| Sibling Difference | 0.13 | -0.02, 0.28 | 0.086 |

**Panel C:**

| | Beta | 95% CI | p-value |
|---|---|---|---|
| GxE | | | |
| Primary Analysis | | | |
| Famine | 0.25 | 0.13, 0.37 | <0.001 |
| PGI | 0.28 | 0.19, 0.36 | <0.001 |
| Famine* PGI | 0.05 | -0.07, 0.17 | 0.452 |
| Exposed vs. Time Control | | | |
| Famine | 0.26 | 0.08, 0.43 | 0.004 |
| PGI | 0.25 | 0.10, 0.40 | 0.002 |
| Famine* PGI | 0.08 | -0.10, 0.26 | 0.380 |

| | | | |
|---|---|---|---|
| Sibling Difference | | | |
| Famine | 0.26 | 0.10, 0.43 | 0.002 |
| PGI | 0.21 | 0.05, 0.38 | 0.012 |
| Famine* PGI | 0.03 | -0.14, 0.19 | 0.758 |

**Table S13. Genetic analysis of in-utero famine exposure effects on Body Mass Index (BMI) adjusted for preconceptual famine exposure (D-1 and D0).** The table shows results from regression analysis of BMI adjusted by exposure to D-1 and D0 (details see Supplemental Table 2). **Panel A** reports analysis of main effects of famine exposure and genetic risk on BMI. Effect-sizes for famine exposure adjusted by D-1 and D0, and genetic risk were estimated in separate models. The "Primary Analysis" model included all participants in our analysis sample with available data on BMI (n=947) and was fitted using linear regression within a generalized estimating equations (GEE) framework to account for non-independence of data from sibling pairs. Models included covariates for D-1, D0, participant age, age-squared, and sex. The "Sibling Difference" model included the subset of the Primary Analysis sample consisting of sibling pairs discordant for famine exposure (n=226 pairs) and was fitted using fixed effects regression. Models included covariates for D-1, D0, age and age-squared (all sibling pairs were of the same sex). The "Exposed vs. Time Control" model included the subset of the Primary Analysis sample consisting of famine exposure participants and their time controls (n= 644) and was fitted using linear regression within a generalized estimating equations (GEE) framework. Effect-sizes are reported as regression beta coefficient and 95% confidence intervals (CI). Genetic association between BMI PGI and BMI were also reported in Panel A. Similar to models assessing main effect of genetic risk which regressed BMI PGI onto BMI, genetic association between BMI PGI and BMI was assessed in three samples: "Famine-exposed" (n = 485), "Time Controls" (n = 161), "Sibling Controls" of famine-exposed (simplified as "Sibling Controls" in the plot legend: n = 226), and "All Controls" consist of time controls, sibling controls of time controls, and sibling controls of famine-exposed (n = 465).

**Panel B** reports analysis of gene-environment correlation (rGE) for BMI. The "Primary Analysis" model included all participants in our analysis sample (n=950) and was fitted using linear regression within a generalized estimating equations (GEE) framework to account for non-independence of data from sibling pairs. Models included covariates for D-1, D0, participant age, age-squared, and sex. The "Sibling Difference" model included the subset of the Primary Analysis sample consisting of sibling pairs discordant for famine exposure (n=226 pairs) and was fitted using linear fixed effects regression. Models included covariates for D-1, D0, age and age-squared (all sibling pairs were of the same sex). The "Exposed vs. Time Control" model included the subset of the Primary Analysis sample consisting of famine exposure participants and their time controls (n= 646) and was fitted using linear regression within a generalized estimating equations (GEE) framework. All rGE effect-sizes were a regression beta coefficient interpretable as Cohen's d (because the PGI is analyzed on standardized scale). **Panel C** reports analysis of gene-environment interaction (GxE) for BMI. Similar as analyses in panel A, a "Primary Analysis" model (n = 934), a "Sibling Difference" model (n=220 pairs), and a "Exposed vs. Time Control" model (n = 632) were constructed with different sub-samples selected from the analysis sample. In each model, coefficients are reported for model terms for famine exposure, genetic risk, and a product term testing their interaction. Coefficients are reported as beta coefficient estimated from linear regression fitted using GEE framework to account for non-independence of sibling data. Models included covariates for D-1, D0, age, age-squared, and sex.

**Body Mass Index**

**Panel A**

| | Beta | 95% CI | p-value |
|---|---|---|---|
| Main Effect of Famine | | | |
| Primary Analysis | 0.28 | 0.16, 0.40 | <0.001 |
| Exposed vs. Time Control | 0.25 | 0.06, 0.43 | 0.009 |
| Sibling Difference | 0.29 | 0.12, 0.46 | <0.001 |
| Main Effect of Famine (adjusted by PGI) | | | |
| Primary Analysis | 0.30 | 0.17, 0.43 | <0.001 |
| Exposed vs. Time Control | 0.31 | 0.13, 0.50 | <0.001 |
| Sibling Difference | 0.30 | 0.12, 0.49 | 0.002 |
| Main Effect of PGI | | | |
| Primary Analysis | 0.31 | 0.24, 0.37 | <0.001 |
| Exposed vs. Time Control | 0.31 | 0.23, 0.38 | <0.001 |
| Sibling Difference | 0.25 | 0.10, 0.40 | <0.001 |
| Genetic Association with BMI | | | |
| Famine-exposed | 0.33 | 0.24, 0.42 | <0.001 |
| Time Controls | 0.23 | 0.09, 0.37 | 0.002 |
| Sibling Controls | 0.25 | 0.13, 0.37 | <0.001 |
| All Controls | 0.28 | 0.19, 0.36 | <0.001 |

**Panel B**

| | Beta | 95% CI | p-value |
|---|---|---|---|
| rGE | | | |
| Primary Analysis | 0.08 | -0.05, 0.20 | 0.217 |
| Exposed vs. Time Control | -0.08 | -0.26, 0.11 | 0.430 |
| Sibling Difference | 0.14 | -0.03, 0.31 | 0.113 |

**Panel C:**

| | Beta | 95% CI | p-value |
|---|---|---|---|
| GxE | | | |
| Primary Analysis | | | |
| Famine | 0.30 | 0.17, 0.43 | <0.001 |
| PGI | 0.28 | 0.19, 0.36 | <0.001 |
| Famine* PGI | 0.04 | -0.10, 0.18 | 0.593 |
| Exposed vs. Time Control | | | |
| Famine | 0.31 | 0.12, 0.49 | 0.001 |
| PGI | 0.25 | 0.10, 0.40 | 0.001 |
| Famine* PGI | 0.06 | -0.12, 0.25 | 0.495 |
| Sibling Difference | | | |

| | | | |
|---|---|---|---|
| Famine | 0.31 | 0.12, 0.49 | 0.002 |
| PGI | 0.20 | 0.04, 0.37 | 0.018 |
| Famine* PGI | 0.07 | -0.13, 0.26 | 0.508 |

**Table S14. Sex stratified genetic analysis, with PRSice2 generated polygenetic index, of in-utero famine exposure effects on Body Mass Index (BMI). Panel A** reports analysis of main effects of famine exposure and genetic risk on BMI. Effect-sizes for famine exposure and genetic risk were estimated in separate models. The "Primary Analysis" model included all participants in our analysis sample with available data on BMI (n=947) and was fitted using linear regression within a generalized estimating equations (GEE) framework to account for non-independence of data from sibling pairs. Models included covariates for participant age, age-squared, and sex. We repeated the main effect analysis in the "Women" (n = 520) and "Men" (n = 427 ) strata separately. Models included covariates for age and age squared. Tests of sex difference were conducted . Coefficients are reported for two product terms: famine and sex, and PGI and sex. Sex interaction coefficients are reported as beta estimates from linear regression fitted using GEE framework to account for non-independence of sibling data. **Panel B** reports analysis of gene-environment correlation (rGE) for BMI polygenic indices. The effect-size is estimated from a linear regression of the BMI PGI on famine exposure fitted using GEE to account for non-independence of data from siblings and including covariates for age, age-squared, and sex. The effect-size is a regression beta coefficient interpretable as Cohen's d (because the PGI is analyzed on standardized scale). We repeated the rGE analysis among women and men separately. Models included covariates for age and age squared. Test of sex difference was conducted. Coefficients are reported for models' interaction term between famine and sex. Sex interaction coefficient is reported as beta estimates from linear regression fitted using GEE framework to account for non-independence of sibling data. **Panel C** reports analysis of gene-environment interaction (GxE) for BMI. Coefficients are reported for model terms for famine exposure, genetic risk, and the product term testing super-multiplicative interaction. Coefficients are reported as beta estimated from linear regression fitted using GEE framework to account for non-independence of sibling data. Models included covariates for age, age-squared, and sex. We repeated the GxE analysis among women and man separately. Models included covariates for age and age squared. Test of sex difference was conducted. Coefficients are reported for three model product terms testing super-multiplicative interaction between famine, PGI, and sex. These interaction coefficients are reported as beta estimates from linear regression fitted using GEE framework to account for non-independence of sibling data.

| Body Mass Index | | | |
|---|---|---|---|
| **Panel A:** | | | |
| | Beta | 95% CI | p-values |
| Main Effect of Famine | | | |
| Primary Analysis | 0.28 | 0.16, 0.40 | <0.001 |
| Women | 0.39 | 0.21, 0.57 | <0.001 |
| Men | 0.14 | -0.01, 0.30 | 0.070 |
| Test of Sex Difference | -0.20 | -0.42, 0.02 | 0.069 |
| Main Effect of PGI | | | |
| Primary Analysis | 0.31 | 0.24, 0.37 | <0.001 |
| Women | 0.36 | 0.27, 0.45 | <0.001 |
| Men | 0.24 | 0.15, 0.33 | <0.001 |
| Test of Sex Difference | -0.12 | -0.25, 0.01 | 0.069 |
| **Panel B:** | | | |
| | Beta | 95% CI | p-value |
| rGE | | | |
| Primary Analysis | 0.09 | -0.03, 0.20 | 0.152 |

| | | | |
|---|---|---|---|
| Women | 0.10 | -0.05, 0.25 | 0.174 |
| Men | 0.05 | -0.13, 0.23 | 0.588 |
| Test of Sex Difference | -0.09 | -0.30, 0.12 | 0.414 |

**Panel C:**

| | Beta | 95%CI | p-value |
|---|---|---|---|
| **GxE** | | | |
| Primary Analysis | | | |
| Famine | 0.25 | 0.13, 0.37 | <0.001 |
| PGI | 0.28 | 0.19, 0.36 | <0.001 |
| Famine:PGI | 0.05 | -0.07, 0.17 | 0.452 |
| Women | | | |
| Famine | 0.35 | 0.18, 0.53 | <0.001 |
| PGI | 0.30 | 0.17, 0.43 | <0.001 |
| Famine:PGI | 0.09 | -0.09, 0.26 | 0.320 |
| Men | | | |
| Famine | 0.24 | 0.14, 0.33 | <0.001 |
| PGI | 0.13 | -0.01, 0.28 | 0.078 |
| Famine:PGI | 0.00 | -0.16, 0.17 | 0.955 |
| Test of Sex Difference | | | |
| Famine:Sex | -0.17 | -0.38, 0.04 | 0.114 |
| PGI:Sex | -0.07 | -0.23, 0.09 | 0.394 |
| Famine:PGI:Sex | -0.08 | -0.32, 0.15 | 0.491 |

**Table S15. Sex stratified genetic analysis adjusted by preconceptual exposure groups (D-1 and D0), with PRSice2 generated polygenetic index, of in-utero famine exposure effects on Body Mass Index (BMI). Panel A** reports analysis of main effects of famine exposure and genetic risk on BMI. Effect-sizes for famine exposure and genetic risk were estimated in separate models. The "Primary Analysis" model included all participants in our analysis sample with available data on BMI (n=947) and was fitted using linear regression within a generalized estimating equations (GEE) framework to account for non-independence of data from sibling pairs. Models included covariates for D-1, D0, participant age, age-squared, and sex. We repeated the main effect analysis in the "Women" (n = 520) and "Men" (n = 427 ) strata separately. Models included covariates for D-1, D0, age and age squared. Tests of sex difference were conducted . Coefficients are reported for two product terms: famine and sex, and PGI and sex. Sex interaction coefficients are reported as beta estimates from linear regression fitted using GEE framework to account for non-independence of sibling data. **Panel B** reports analysis of gene-environment correlation (rGE) for BMI polygenic indices. The effect-size is estimated from a linear regression of the BMI PGI on famine exposure fitted using GEE to account for non-independence of data from siblings and including covariates for D-1, D0, age, age-squared, and sex. The effect-size is a regression beta coefficient interpretable as Cohen's d (because the PGI is analyzed on standardized scale). We repeated the rGE analysis among women and men separately. Models included covariates for D-1, D0, age and age squared. Test of sex difference was conducted. Coefficients are reported for models' interaction term between famine and sex. Sex interaction coefficient is reported as beta estimates from linear regression fitted using GEE framework to account for non-independence of sibling data. **Panel C** reports analysis of gene-environment interaction (GxE) for BMI. Coefficients are reported for model terms for famine exposure, genetic risk, and the product term testing super-multiplicative interaction. Coefficients are reported as beta estimated from linear regression fitted using GEE framework to account for non-independence of sibling data. Models included covariates for D-1, D0, age, age-squared, and sex. We repeated the GxE analysis among women and man separately. Models included covariates for age and age squared. Test of sex difference was conducted. Coefficients are reported for three model product terms testing super-multiplicative interaction between famine, PGI, and sex. These interaction coefficients are reported as beta estimates from linear regression fitted using GEE framework to account for non-independence of sibling data.

| | Body Mass Index | | |
|---|---|---|---|
| **Panel A:** | | | |
| | Beta | 95% CI | p-values |
| Main Effect of Famine | | | |
| Primary Analysis | 0.32 | 0.19, 0.46 | <0.001 |
| Women | 0.50 | 0.30, 0.70 | <0.001 |
| Men | 0.12 | -0.05, 0.28 | 0.172 |
| Test of Sex Difference | -0.20 | -0.42, 0.02 | 0.069 |
| Main Effect of PGI | | | |
| Primary Analysis | 0.31 | 0.24, 0.37 | <0.001 |
| Women | 0.36 | 0.27, 0.45 | <0.001 |
| Men | 0.24 | 0.15, 0.33 | <0.001 |
| Test of Sex Difference | -0.12 | -0.25, 0.01 | 0.069 |

| **Panel B:** | | | |
|---|---|---|---|
| | Beta | 95% CI | p-value |
| rGE | | | |

| | | | |
|---|---|---|---|
| Primary Analysis | 0.08 | -0.05, 0.20 | 0.217 |
| Women | 0.07 | -0.09, 0.24 | 0.371 |
| Men | 0.05 | -0.14, 0.24 | 0.578 |
| Test of Sex Difference | -0.09 | -0.30, 0.12 | 0.414 |

**Panel C:**

| | Beta | 95%CI | p-value |
|---|---|---|---|
| **GxE** | | | |
| Primary Analysis | | | |
| Famine | 0.30 | 0.17, 0.43 | <0.001 |
| PGI | 0.28 | 0.19, 0.36 | <0.001 |
| Famine:PGI | 0.04 | -0.10, 0.18 | 0.593 |
| Women | | | |
| Famine | 0.47 | 0.28, 0.67 | <0.001 |
| PGI | 0.30 | 0.17, 0.43 | <0.001 |
| Famine:PGI | 0.09 | -0.11, 0.29 | 0.369 |
| Men | | | |
| Famine | 0.11 | -0.05, 0.26 | 0.184 |
| PGI | 0.23 | 0.14, 0.33 | <0.001 |
| Famine:PGI | -0.02 | -0.21, 0.17 | 0.864 |
| Test of Sex Difference | | | |
| Famine:Sex | -0.17 | -0.38, 0.04 | 0.114 |
| PGI:Sex | -0.07 | -0.23, 0.09 | 0.394 |
| Famine:PGI:Sex | -0.08 | -0.32, 0.15 | 0.491 |

**Table S16. Sensitivity analysis on the timing-specific gene-environment correlation between in-utero famine exposure and PRSice2 produced body mass index (BMI) polygenic index (PGI).** This table report the analysis of timing-specific gene-environment correlation between famine exposure and genetic risk on BMI. Effect-sizes for six famine exposure groups (D4 ~ D-1) were estimated in a model which included all participants in our analysis sample with available data on BMI (n=950) and was fitted using linear regression within a generalized estimating equations (GEE) framework to account for non-independence of data from sibling pairs. Model regressed genetic risk of elevated BMI onto six famine exposure groups and adjusted for covariates of participant age, age-squared, and sex.

| Timing-specific gene-environment correlation | | | |
|---|---|---|---|
| | Beta | 95% CI | p-values |
| D4 (weeks 31 to delivery) | 0.05 | -0.13, 0.24 | 0.585 |
| D3 (gestational weeks 21-30) | 0.14 | -0.03, 0.31 | 0.110 |
| D2 (gestational weeks 11-20) | -0.10 | -0.29, 0.08 | 0.289 |
| D1 (gestational weeks 1-10) | 0.19 | -0.03, 0.41 | 0.086 |
| D0 (conception to less than 10 weeks) | 0.02 | -0.21, 0.26 | 0.849 |
| D-1 (preconceptional) | 0.15 | -0.08, 0.38 | 0.196 |

**Figure S3. Sensitivity analysis on the timing-specific gene-environment correlation between in-utero famine exposure and PRSice2 produced body mass index polygenic indices.** This plot shows the correlation between genetic risk and timing-specific famine exposures. Timing of famine exposure was shown on the x-axis. The beta effect sizes estimated from linear regression of the BMI PGI on famine exposure with six time-specific exposure windows added as covariates were shown on the y-axis . Model was fitted using generalized estimating equations (GEE) framework to account for non-independence of data from sibling pairs.

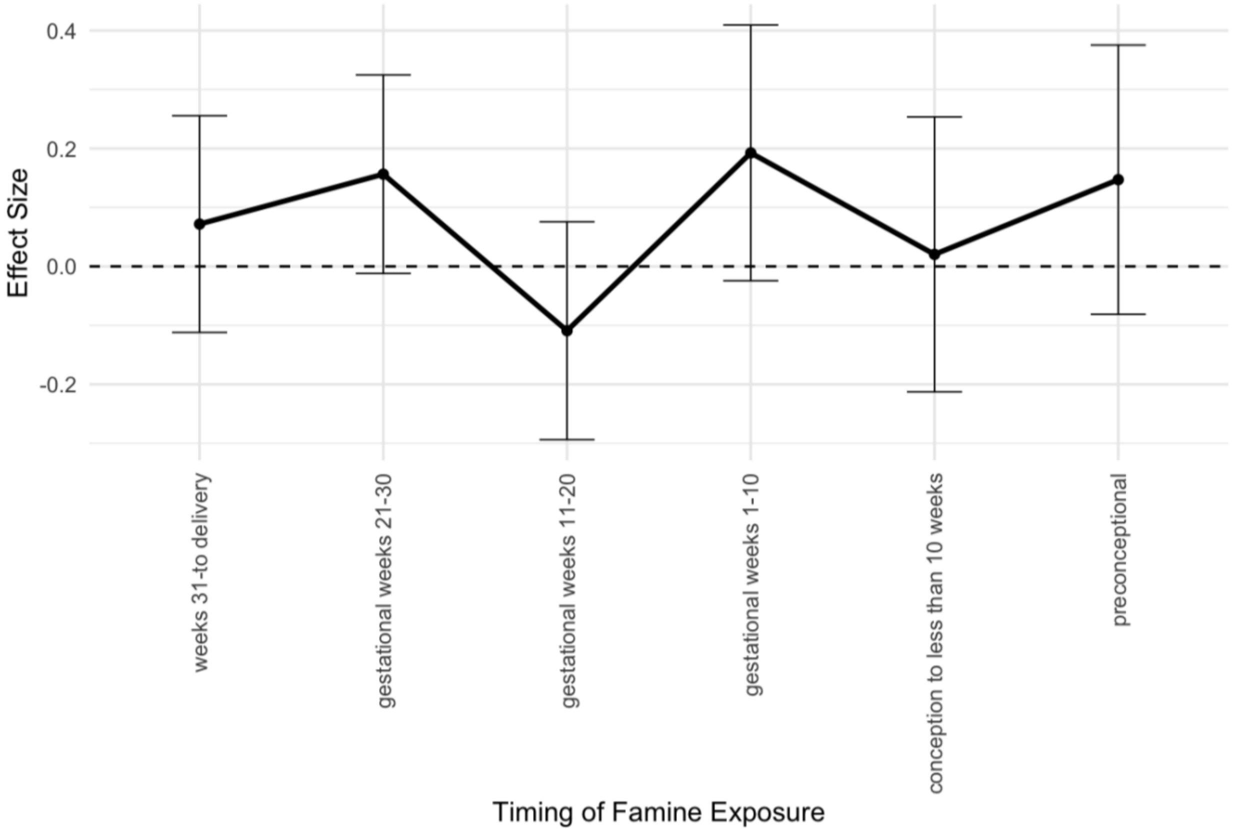

**Table S17. Sensitivity analysis on the timing-specific gene-environment interaction between in-utero famine exposure and PRSice2 produced body mass index (BMI) polygenic indices (PGI).** This table report the analysis of timing-specific gene-environment interaction between famine exposure and genetic risk on BMI. Effect-sizes for six product terms between famine exposure groups (D4 ~ D-1)and genetic risk of elevated BMI were estimated in a model which included all participants in our analysis sample with available data on BMI (n=950) and was fitted using linear regression within a generalized estimating equations (GEE) framework to account for non-independence of data from sibling pairs. Model regressed BMI onto six product terms between famine exposure groups and genetic risk of elevated BMI. Model was adjusted for covariates of participant age, age-squared, and sex.

| | Timing-specific gene-environment interaction | | |
|---|---|---|---|
| | Beta | 95% CI | p-values |
| PGI:D4 (weeks 31 to delivery) | 0.19 | 0.00, 0.39 | 0.051 |
| PGI:D3 (gestational weeks 21-30) | 0.19 | -0.03, 0.41 | 0.093 |
| PGI:D2 (gestational weeks 11-20) | 0.13 | -0.08, 0.34 | 0.224 |
| PGI:D1 (gestational weeks 1-10) | 0.39 | 0.14, 0.65 | 0.003 |
| PGI:D0 (conception to less than 10 weeks) | 0.41 | 0.25, 0.57 | <0.001 |
| PGI:D-1 (preconceptional) | -0.05 | -0.27, 0.17 | 0.664 |

**Figure S4. Sensitivity analysis on the timing-specific gene-environment interaction between in-utero famine exposure and PRSice2 produced body mass index (BMI) polygenic indices.** This plot shows the interaction between genetic risk and timing-specific famine exposures. Timing of famine exposure was shown on the x-axis. The beta coefficients representing the genetic effect in the famine exposed group were shown on the y-axis. These beta coefficients were estimated from linear regression of the BMI PGI on famine exposure with six product terms between PGI and the exposure windows added as covariates. Model was fitted using generalized estimating equations (GEE) framework to account for non-independence of data from sibling pairs. The grey line is showing the reference main effect of genetic risk for BMI without adjusting for timing-specific covariates, which was obtained in the primary main genetic effect analysis.

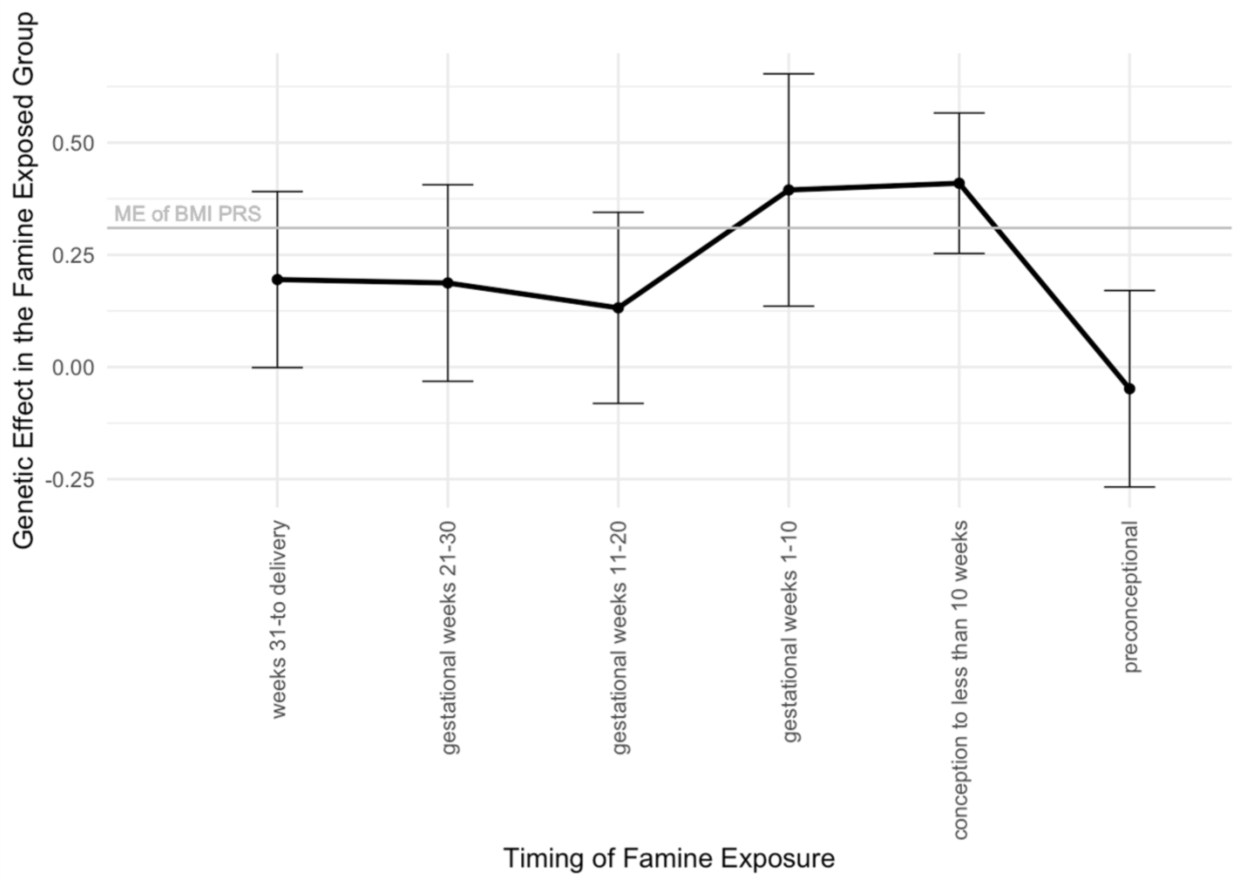
